# Positive mental health among children 11 years and under in Western countries: a scoping review to inform Canada’s public health surveillance

**DOI:** 10.1101/2025.10.08.25337090

**Authors:** Melanie Varin, Mihojana Jhumi, Colin A. Capaldi, Raelyne Dopko, Laura L. Ooi

## Abstract

**Background:** The Public Health Agency of Canada developed the Positive Mental Health Surveillance Indicator Framework (PMHSIF), which is used to monitor positive mental health (PMH) and its determinants in Canada. While adult and youth versions of the PMHSIF were released, additional research is needed to identify relevant and age-appropriate concepts of PMH among children.

**Methods:** A scoping review of peer-reviewed and grey literature was conducted to examine how PMH is conceptualized and measured among children (< 12 years). Online academic databases were searched up until January 31, 2023. Grey literature sources were conducted up until October 3, 2023. Data on study characteristics were extracted and some were tallied and explained. Measures of PMH were categorized as self- or other-rated.

**Results:** A total of 636 documents were identified through the search strategies. Of these, 65 documents from the grey literature and 39 peer-reviewed papers were included in this review. Many of the articles (74%) mentioned at least one theory in the introduction/background. The PMH concepts that emerged included: hedonic well-being (n=77), psychological well-being (n=41), social well-being (n=30), and social emotional learning and/or positive development (n=19). Various risk and protective factors were extracted.

**Conclusion:** The findings support the use of the existing Canadian PMH conceptual framework for children, albeit with small modifications to ensure that child-relevant pieces are reflected. Additional work is needed before these results can be used for national surveillance. This review addresses a gap in the literature, encourages routine reporting of child PMH across Canada, and better informs public health policy.

## Background

Much attention has been dedicated to measuring and monitoring well-being during childhood as it is a developmental period that can impact future social, emotional, cognitive and health outcomes (Adamson P, 2013; Helseth & Haraldstad, 2014; Lippman LH et al., 2011; OECD, 2021; OECD, 2022; UNICEF, 2020). The conceptualization of well-being is often broad and can differ based on discipline (e.g., psychology, philosophy, public policy, economics) (Jarden & Roache, 2023; OECD, 2024; UNICEF, 2020; UNICEF Canada, 2019). There is no definition of well-being that is universally agreed upon (Soffia & Turner, 2021); however, it is often used synonymously with quality of life, and can include objective (e.g., income, education, life expectancy) and subjective (e.g., life satisfaction) indicators (Department of Finance Canada, 2021; Statham & Chase, 2010). While there has been substantial work on well-being among children (Adamson P, 2013; Lippman LH et al., 2011; Moore KA et al., 2012; OECD, 2022; UNICEF, 2020), the focus has mainly been on objective measures with considerable gaps on subjective aspects such as child mental well-being, which can also be referred to as positive mental health (PMH) (Marquez J et al., 2024; OECD, 2024; Soffia & Turner, 2021; UNICEF, 2020).

The Public Health Agency of Canada defines PMH as “the capacity of each and all of us to feel, think, [and] act in ways that enhance our ability to enjoy life and deal with the challenges we face” (PHAC, 2014). It includes feeling good (hedonic or emotional well-being) and functioning well (eudaimonic or psychological and social well-being) (Orpana et al., 2016). Many member countries of the Organisation for Economic Co-operation and Development (OECD), including Canada, report on national indicators of PMH (OECD, 2023; PHAC, 2024; Ruggeri et al., 2020). Since 2016, the Public Health Agency of Canada has been conducting national surveillance of PMH through its Positive Mental Health Surveillance Indicator Framework (PMHSIF), which is based on a conceptual socioecological model that highlights important risk and protective factors across four different domains (individual, family, community, societal) (Orpana et al., 2016). Currently, five indicator groups of PMH outcomes (self-rated mental health, life satisfaction, happiness, psychological well-being, and social well-being) and related risk and protective factors are monitored nationally for youth (ages 12-17 years) and adults (ages 18 years and over) (PHAC, 2024). However, there is no routine reporting at the national level for children (11 years and under). As such, less is known about PMH among this population (Hepburn & Daneman, 2015; OECD, 2024). There is a growing need to identify what PMH concepts are important for children and how to measure them (OECD, 2021; OECD, 2024). Monitoring PMH at the national level among children in Canada is important to understand how they are feeling and functioning, which could help inform policies aimed at improving current and future PMH (OECD, 2021). Furthermore, PMH in childhood can influence development, academic performance (Durlak et al., 2011), physical health, social health, and future mental health (Canny M et al., 2017; Public Health England, 2015), which provides additional support for the importance of examining and reporting on this outcome.

The conceptualization and measurement of PMH as well as the associated risk and protective factors during childhood should reflect the perspectives, experiences, and needs of children, which may differ from those of youth and adults. Although the conceptual framework and overall structure of Canada’s PMHSIF (Orpana et al., 2016) may also be appropriate for children, specific indicators and/or measures for youth and adults are likely less relevant for children (e.g., substance use) and potentially important child-specific indicators may be missing. Indeed, the identification of indicators and measures included in Canada’s PMHSIF was largely based on evidence from adults and youth (Orpana et al., 2016), which signals a need to review the literature with a child-specific focus. There are additional layers of complexity when measuring constructs for children that need to be considered. For example, measures that have been developed for children are frequently other-rated (e.g., by a parent or a teacher), but discrepancies have been observed between child self-reported and other-reported measures of child mental health (Achenbach et al., 1987; Caqueo-Urízar et al., 2022; De Los Reyes et al., 2015; Ederer EM, 2004; Holte et al., 2014; Van Roy et al., 2010). Children may also have less experience or capacity to report on some of their internal states compared to youth and adults (e.g., they may not fully understand the concept they are being asked about if proper methodologies are not implemented and/or developmental factors are not taken into consideration) (Spieth LE, 2001).

Thus, to appraise the potential for the development of a child PMHSIF and better understand the applicability of the existing PMH conceptual framework (Orpana et al., 2016) and the PMHSIFs for youth and adults (PHAC, 2024) to children’s PMH, an extensive review of both peer-reviewed and grey literature specific to child (aged 11 years and under) PMH was conducted to answer the following questions:

1. What are the theoretical and conceptual underpinnings identified for child PMH?
2. What are PMH concepts/outcomes for children and are there instruments to measure them?
3. What are the PMH determinants (risk and protective factors) among children?

## Methods

A scoping review of peer-reviewed and grey literature on PMH among children was conducted in accordance with the PRISMA-ScR (PRISMA extension for Scoping Reviews) (Tricco et al., 2018). There is no publicly available registered protocol for this review.

### Eligibility criteria

The purpose of this scoping review was to appraise the potential for the development of a PMHSIF for children aged 11 years and under in Canada. As such, the scoping review was restricted to that age range and to Western countries for comparability purposes. If the age of the sample was ambiguous during initial title and abstract screening (e.g., only a mean age was provided in the abstract and it could not be confirmed that children ≤ 11 years were included), the study was screened in for a methods-specific review. If there were no children ≤ 11 years or if no information other than a mean age greater than 11 years was provided (e.g., there was no age range), the study was excluded. Studies where the age range of the population was specified were reviewed and the ones with an age range that overlapped between children and youth were screened in to maximize the amount of information we had for children (even if it included some youth). In instances where no study location was provided, we used the first author’s affiliation. Certain types of evidence were excluded (e.g., experimental studies, theses/dissertations) to align with the methodology used in a previous review conducted during the development of the PMHSIF (Orpana et al., 2016). The current scoping review was restricted to studies that included information on PMH (e.g., emotional, psychological, or social well-being). Studies solely examining indicators of ‘negative’ mental health (e.g., mental illness, mental health services and utilization) were excluded as they were out of scope. To maximize the amount of evidence, we did not restrict the publication start date. Lastly, studies were restricted to those written in English or French (i.e., the two official languages in Canada). The full list of inclusion and exclusion criteria can be found in Table 1.

**Table 1.** Inclusion and exclusion criteria.

| Category | Inclusion | Exclusion |
| --- | --- | --- |
| Population of interest | Children 11 years of age and under (can be any age group within that range) | Studies with populations solely 12 years and above |
| Study design/types of evidence | *Peer-reviewed:* Studies, reviews (narrative literature reviews, scoping reviews, systematic reviews), and meta-analyses<br>*Grey:* Non peer-reviewed sources that report empirical evidence, government reports, research reports, position papers, webpages, non-governmental sources, infographics, data blogs, and book chapters<br>*Both:* Quantitative and qualitative evidence, and theoretical/conceptual papers | *Peer-reviewed:* Conference abstracts, editorials, case reports, case series, experimental studies, full books, and theses/dissertations<br>*Grey:* News articles and multimedia sources (such as podcasts) |
| Content | Emotional, psychological, or social well-being from a public health, PMH, or mental health promotion perspective | Mental health or well-being concept that does not include at least one aspect specific to PMH<br>Focus is on ‘negative’ mental health<br>Concept does not fall under or cannot be linked to the Public Health Agency of Canada’s definition of PMH<br>PMH outcome is in adulthood (even if determinants are in childhood) |
| Country | *Western countries:* European Union, United Kingdom, Norway, Iceland, Switzerland, the Americas (North and South), Australia, and New Zealand | All other countries not listed in left column |
| Publication date | *Peer-reviewed:* Database inception to January 31, 2023<br>*Grey:* Inception to October 3, 2023 |  |
| Publication language | French or English | Sources that were not published in French or English |

### Information sources and search strategy

The search strategies were developed through collaboration with a research librarian. The librarian completed a peer-reviewed literature search through the electronic databases Ovid MEDLINE, Embase, PsycINFO, and SCOPUS up until January 31, 2023. The complete search strategy for peer-reviewed articles can be found in Appendix 1. For the grey literature review, the same librarian provided a set of search strategies to complete general searches using Google and site-specific searches, as well as direct links to specific sources to screen. Grey literature sources were searched until October 3, 2023. A complete list of grey literature sources and search strings can be found in Appendix 2.

### Screening and data extraction

For the peer-reviewed literature, title and abstract screening were conducted by three reviewers (MV, LLO, FLP). When disagreement in selection choices occurred in the title and abstract screening, the reviewers discussed their reasoning to come to a consensus. Full-text screening was done by two reviewers (MV, FLP). Data extraction was conducted by a separate analyst (MJ). The following data elements were extracted: title, author(s), year of publication, type of paper (conceptual or empirical), location (where the study was conducted), aims/purpose, summary of introduction including theoretical/conceptual underpinning(s), age/developmental stage, sample size, PMH outcome/concept(s), instrument(s) capturing PMH concept(s), PMH risk and/or protective factor(s), key finding(s), and additional information (e.g., if there were more findings but we only extracted information specific to PMH, relevant appendices).

For the grey literature, two epidemiologists (LLO, MV) screened the sources based on the inclusion criteria. The project lead (MV) independently extracted the following data elements: title, location (country, province/state), year of publication, summary of background including theoretical/conceptual underpinning(s), age/developmental stage, sample size (if applicable), PMH outcome/concept(s), instrument(s) capturing PMH concept(s), PMH risk and/or protective factor(s), key finding(s), and additional information.

### Synthesis of results

For the sources included in the scoping review, the following characteristics were tallied overall and by type of source (i.e., grey and peer-reviewed): publication date, source location, type of evidence, age/developmental stage, and language. The theory and conceptual underpinnings that were extracted from the introduction/background section for each source were tallied and a brief explanation was provided for each. Overall concepts that encompassed different PMH outcomes were identified, tallied, and explained. Instruments used to measure PMH outcomes were categorized into one of two categories: self-rated or other-rated. Lastly, to align with the existing PMHSIF structure (Orpana et al., 2016), high-level determinants with examples were categorized into the following four socio-ecological domains: individual, family, community, and society.

## Results

A total of 636 documents (448 from the peer-reviewed databases and 188 from grey literature sources) were screened through this scoping review’s search strategy. After screening, a total of 104 sources (39 from the peer-reviewed literature and 65 from the grey literature) were included in this scoping review. The PRISMA-Scr flow chart can be found in Figure 1. The list of all 104 sources included in the scoping review (along with their source ID) can be found in Table 2.

**Figure 1.**
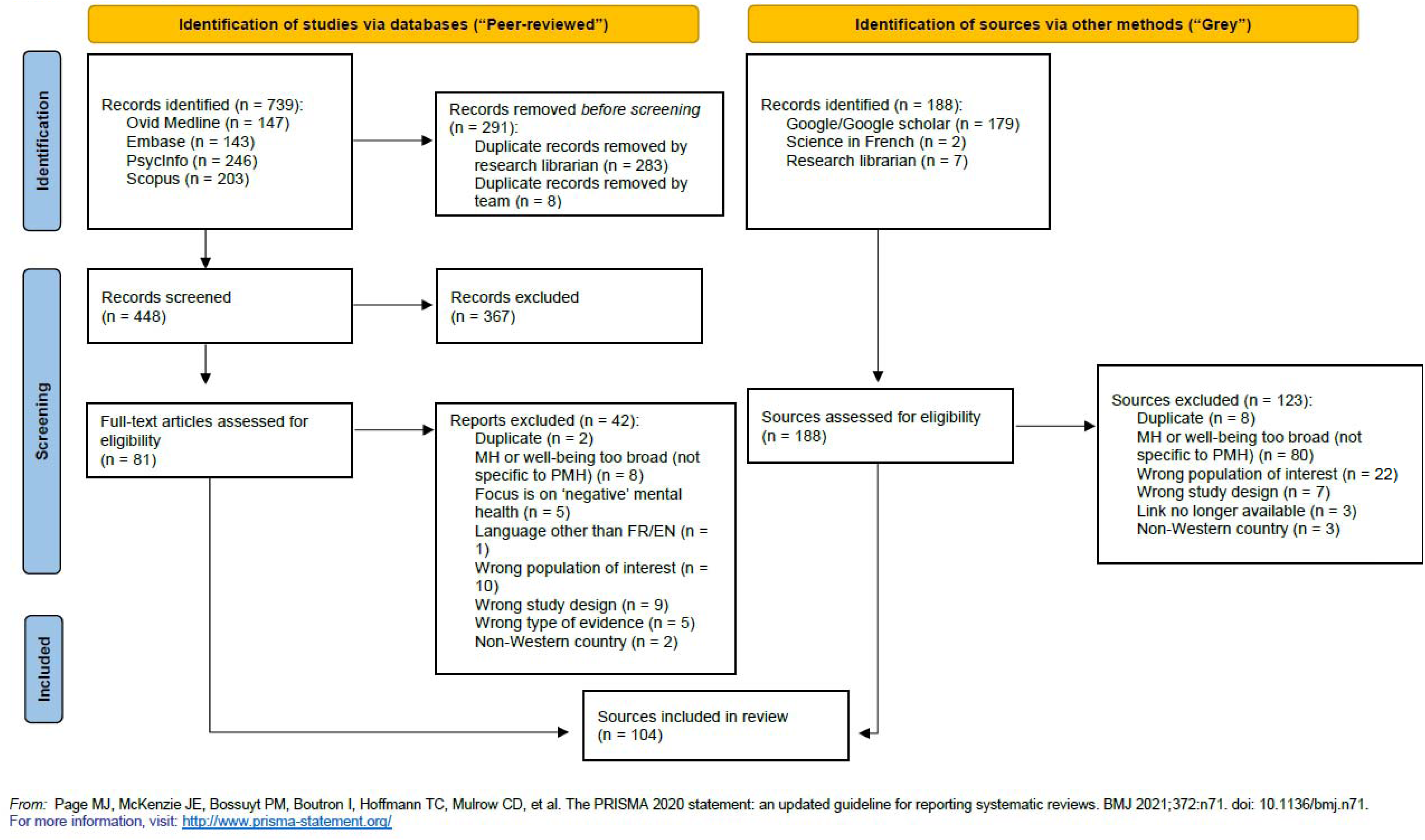
Flow chart. **Abbreviations**: MH, Mental health; PMH, Positive mental health; FR/EN, French/English

**Table 2.** List of sources included in scoping review with source ID.

| Source ID | Reference |
| --- | --- |
| 1 | Cho, E.Y.-N.; Yu, F.-Y. A review of measurement tools for child wellbeing. *Children and Youth Services Review* 2020;119. https://doi.org/10.1016/j.childyouth.2020.105576. |
| 2 | Nagy B.E, Kovács K.E, Varga T, Boris P, Józsa T, Oláh R. A review on children and adolescents' mental health indicators: Towards a valid mental health assessment in Hungarian youth *Cognition, Brain, Behavior. An Interdisciplinary Journal* 2021;25(4). https://dx.doi.org/10.24193/cbb.2021.25.18. |
| 3 | Lawlor, M.S., Schonert-Reichl, K.A., Gadermann, A.M. et al. A Validation Study of the Mindful Attention Awareness Scale Adapted for Children. *Mindfulness* 5, 730–741 (2014). https://doi.org/10.1007/s12671-013-0228-4. |
| 4 | Neubauer AB, Kramer AC, Schmiedek F. Assessing domain-general need fulfillment in children and adults: Introducing the General Need Satisfaction and Frustration scale. *Psychol Assess.* 2022 Nov;34(11):1022-1035. https://doi.org/10.1037/pas0001169. |
| 5 | Hope S, Rougeaux E, Deighton J, Law C, Pearce A. Associations between mental health competence and indicators of physical health and cognitive development in eleven year olds: findings from the UK Millennium Cohort Study. *BMC Public Health.* 2019 Nov 6;19(1):1461. https://doi.org/10.1186/s12889-019-7789-7. |
| 6 | González-Carrasco, M., Casas, F., Malo, S. et al. Changes with Age in Subjective Well-Being Through the Adolescent Years: Differences by Gender. *J Happiness Stud* 18, 63–88 (2017). https://doi.org/10.1007/s10902-016-9717-1. |
| 7 | Huebner, E.S., Suldo, S.M., Valois, R.F. (2005). Children's Life Satisfaction. In: Moore, K.A., Lippman, L.H. (eds) *What Do Children Need to Flourish?. The Search Institute Series on Developmentally Attentive Community and Society*, vol 3. Springer, Boston, MA . https://doi.org/10.1007/0-387-23823-9_4. |
| 8 | Moore, S.A., Widaes-Benitez, O., Carnazzo, K.W. et al. Conducting Universal Complete Mental Health Screening via Student Self-Report. *Contemp School Psychol* 19, 253–267 (2015). https://doi.org/10.1007/s40688-015-0062-x. |
| 9 | Vaqué-Crusellas, C., González, M. & Casas, F. Does Satisfaction with Food Matter? Testing the Personal Well-Being Index-School Children (PWI-SC) with an Additional Item on Satisfaction with Food on a Sample of 10 to 12-Year-Olds. *Child Ind Res* 8, 961–973 (2015). https://doi.org/10.1007/s12187-015-9301-y. |
| 10 | Lawler, M.J., Newland, L.A., Giger, J.T. et al. Ecological, Relationship-Based Model of Children's Subjective Well-Being: Perspectives of 10-Year-Old Children in the United States and 10 Other Countries. *Child Ind Res* 10, 1–18 (2017). https://doi.org/10.1007/s12187-016-9376-00. |
| 11 | Casas, F., González-Carrasco, M., Oriol, X. et al. Economic and Children's Subjective Well-Being Indicators at the National Level in 35 Countries. *Child Ind Res* 15, 1539–1563 (2022). https://doi.org/10.1007/s12187-022-09918-4. |
| 12 | Mériaux BG, Berg M, Hellström AL. Everyday experiences of life, body and well-being in children with overweight. *Scand J Caring Sci.* 2010 Mar;24(1):14-23. https://doi.org/10.1111/j.1471-6712.2008.00678.x. |
| 13 | Allen K, Marlow R, Edwards V, Parker C, Rodgers L, Ukoumunne OC, Seem EC, Hayes R, Price A, Ford T. 'How I Feel About My School': The construction and validation of a measure of wellbeing at school for primary school children. *Clin Child Psychol Psychiatry*. |
|  | 2018 Jan;23(1):25-41. https://doi.org/10.1177/1359104516687612. |
| 14 | Moore, K. A., Lippman, L., & Brown, B. (2004). Indicators of Child Well-Being: the Promise for Positive Youth Development. The ANNALS of the American Academy of Political and Social Science, 591(1), 125-145. https://doi.org/10.1177/0002716203260103. |
| 15 | Moore, K.A., Murphey, D., Bandy, T., Lawner, E. (2014). Indices of Child Well-Being and Developmental Contexts. In: Ben-Arieh, A., Casas, F., Frønes, I., Korbin, J. (eds) Handbook of Child Well-Being. Springer, Dordrecht. https://doi.org/10.1007/978-90-481-9063-8_139. |
| 16 | Rountree J, Smith A. Strength-based well-being indicators for Indigenous children and families: A literature review of Indigenous communities' identified well-being indicators. Am Indian Alsk Native Ment Health Res. 2016;23(3):206-20. https://doi.org/10.5820/aian.2303.2016.206. |
| 17 | Moula Z, Walshe N, Lee E. Making Nature Explicit in Children's Drawings of Wellbeing and Happy Spaces. Child Indic Res. 2021;14(4):1653-1675. https://doi.org/10.1007/s12187-021-09811-6. |
| 18 | Casas, F., Rees, G. Measures of Children's Subjective Well-Being: Analysis of the Potential for Cross-National Comparisons. Child Ind Res 8, 49–69 (2015). https://doi.org/10.1007/s12187-014-9293-z. |
| 19 | Crouch SR, Waters E, McNair R, Power J, Davis E. Parent-reported measures of child health and wellbeing in same-sex parent families: a cross-sectional survey. BMC Public Health. 2014 Jun 21;14:635. https://doi.org/10.1186/1471-2458-14-635. |
| 20 | Scherr, S., Mares, M.-L., Bartsch, A., & Goetz, M. (2019). Parents, Television, and Children's Emotional Expressions: A Cross-Cultural Multilevel Model. Journal of Cross-Cultural Psychology, 50(1), 22-46. https://doi.org/10.1177/0022022118806585. |
| 21 | Lawrence, J.A., Kaplan, I. & Collard, A.H. Perspectives of Refugee Children Resettling in Australia on Indicators of Their Wellbeing. Child Ind Res 12, 943–962 (2019). https://doi.org/10.1007/s12187-018-9568-x. |
| 22 | Moore KA, Murphey D, Bandy T. Positive child well-being: an index based on data for individual children. Matern Child Health J. 2012 Apr;16 Suppl 1:S119-28. https://doi.org/10.1007/s10995-012-1001-3. |
| 23 | Lippman, L.H., Moore, K.A. & McIntosh, H. Positive Indicators of Child Well-Being: A Conceptual Framework, Measures, and Methodological Issues. Applied Research Quality Life 6, 425–449 (2011). https://doi.org/10.1007/s11482-011-9138-6. |
| 24 | Newland, L.A., Lawler, M.J., Giger, J.T. et al. Predictors of Children's Subjective Well-Being in Rural Communities of the United States. Child Ind Res 8, 177–198 (2015). https://doi.org/10.1007/s12187-014-9287-x. |
| 25 | Goldfeld S, Kvalsvig A, Incledon E, O'Connor M, Mensah F. Predictors of mental health competence in a population cohort of Australian children. J Epidemiol Community Health. 2014 May;68(5):431-7. https://doi.org/10.1136/jech-2013-203007. |
| 26 | O'Connor, E., O'Connor, M., Gray, S. et al. Profiles of Mental Health Competence and Difficulties as Predictors of Children's Early Learning. School Mental Health 10, 402–416 (2018). https://doi.org/10.1007/s12310-018-9252-9. |
| 27 | O'Brien, W., Belton, S., Fitzpatrick, B., Shannon, S., Brennan, D., Chambers, F., ... Breslin, G. (2021). Relationship between Gender, Physical Activity, Screen Time, Body Mass Index and Wellbeing in Irish Children from Social-Disadvantage. Child Care in Practice, 30(3), 220–234. https://doi.org/10.1080/13575279.2021.1887815. |
| 28 | Denham SA, Kalb S, Way E, Warren-Khot H, Rhoades BL, Bassett HH. Social and emotional information processing in preschoolers: Indicator of early school success? Early Child Dev Care. 2013 Jan 1;183(5):667-688. https://doi.org/10.1080/03004430.2012.682728. |
| 29 | Abrahams L, Pancorbo G, Primi R, Santos D, Kyllonen P, John OP, De Fruyt F. Social-emotional skill assessment in children and |
|  | adolescents: Advances and challenges in personality, clinical, and educational contexts. Psychol Assess. 2019 Apr;31(4):460-473. https://doi.org/10.1037/pas0000591. |
| 30 | Ravens-Sieberer U, Devine J, Bevens K, Riley AW, Moon J, Salsman JM, Forrest CB. Subjective well-being measures for children were developed within the PROMIS project: presentation of first results. J Clin Epidemiol. 2014 Feb;67(2):207-18. https://doi.org/10.1016/j.jclinepi.2013.08.018. |
| 31 | Moore, K. A., & Glei, D. (1995). Taking the Plunge: An Examination of Positive Youth Development. Journal of Adolescent Research, 10(1), 15-40. https://doi.org/10.1177/0743554895101003. |
| 32 | DuBois, D. L., Felner, R. D., Lockerd, E. M., Parra, G. R., & Lopez, C. (2003). The quadripartite model revisited: Promoting positive mental health in children and adolescents. In M. A. Reinecke, F. M. Dattilio, & A. Freeman (Eds.), Cognitive therapy with children and adolescents: A casebook for clinical practice (2nd ed., pp. 402–433). The Guilford Press. |
| 33 | Caputi, M., & Bosacki, S. (2023). The Role of Gender in the Relation Among Anxiety, Theory of Mind, and Well-Being in Early Adolescents. The Journal of Early Adolescence, 43(7), 845-866. https://doi.org/10.1177/02724316221130440. |
| 34 | Gross-Manos, D. (2014). The Role of Peers in Children's Lives and Their Contribution to Child Well-Being: Theory and Research. In: Ben-Arieh, A., Casas, F., Frønes, I., Korbin, J. (eds) Handbook of Child Well-Being. Springer, Dordrecht. https://doi.org/10.1007/978-90-481-9063-8_176. |
| 35 | Savahl S, Casas F, Adams S. The Structure of Children's Subjective Well-being. Front Psychol. 2021 Jun 11;12:650691. https://doi.org/10.3389/fpsyg.2021.650691. |
| 36 | Castellá Sarriera, J., & Bedin, L. M. (2015). Towards a socio-community model: A well-being approach. Universitas Psychologica, 14(4), 1387–1398. https://doi.org/10.11144/Javeriana.upsy14-4.tscm. |
| 37 | Smees R, Rinaldi LJ, Simner J. Well-being measures for younger children. Psychol Assess. 2020 Feb;32(2):154-169. https://doi.org/10.1037/pas0000768. |
| 38 | Gordon, G. (2014). Well played: The origins and future of playfulness. American Journal of Play, 6(2), 234–266. |
| 39 | Ash, C., & Huebner, E. S. (1998). Life satisfaction reports of gifted middle-school children. School Psychology Quarterly, 13(4), 310–321. https://doi.org/10.1037/h0088987. |
| 40 | Centers for Disease Control and Prevention. Data and Statistics on Children's Mental Health. US: CDC; 2023. Available from: https://www.cdc.gov/childrensmentalhealth/data.html. |
| 41 | Public Health Scotland. Children and young people's mental health indicators. UK; 2012. Available from: https://www.healthscotland.scot/health-topics/mental-health-and-wellbeing/children-and-young-peoples-mental-health-indicators. |
| 42 | Public Health England. Children and Young People's Mental Health and Wellbeing. UK; 2016. Available from: https://assets.publishing.service.gov.uk/media/5a80c3e240f0b62305b8d06c/Mental_health_of_children_in_England.pdf. |
| 43 | Public Health England. Measuring mental wellbeing in children and young people. UK; 2015. Available from: https://assets.publishing.service.gov.uk/media/5c2f66daed915d731281fdc2/Measuring_mental_wellbeing_in_children_and_young_people.pdf. |
| 44 | Child Trends. Common Indicators of Social-Emotional Well-being in Early Childhood. USA; 2017. Available from: https://www.childtrends.org/project/common-indicators-of-social-emotional-well-being-in-early-childhood. |
| 45 | Office for National Statistics. Children's well-being indicator review, UK; 2020. UK; 2020. Available from: https://www.ons.gov.uk/peoplepopulationandcommunity/wellbeing/articles/childrenswellbeingindicatorreviewuk2020/2020-09-02#proposed-ons-childrens-well-being-indicator-set. |
| 46 | Australian Institute of Health and Welfare. Australia's children: social and emotional wellbeing. Australia; 2020. Available from: https://www.aihw.gov.au/reports/children-youth/australias-children/contents/health/social-emotional-wellbeing. |
| 47 | Schonert-Reichl KA, Lawlor MA, Oberle E, Thomson K. Identifying indicators and tools for measuring social and emotional healthy living: children ages 5-12. Canada: University of British Columbia; 2009. Available from: http://www.jcsh-cces.ca/upload/SEL%20Report%20FINAL_May26.pdf. |
| 48 | Australian Government Department of Social Services. Family and Children Activity outcomes framework. Australia; 2021. Available from: https://www.dss.gov.au/sites/default/files/documents/12_2021/families-and-children-activity-outcomes-framework-suggested-indicators.pdf. |
| 49 | What Works Centre for Wellbeing. Measuring Children and Young People's Subjective Wellbeing: A conceptual framework. UK; 2021. Available from: https://whatworkswellbeing.org/wp-content/uploads/2021/10/MCYPSW-Conceptual-framework-2.pdf. |
| 50 | Tillmann S, Tobin D, Avison W, Gilliland J. Mental health benefits of interactions with nature in children and teenagers: a systematic review. J Epidemiol Community Health. 2018 Oct;72(10):958-966. https://doi.org/10.1136/jech-2018-210436. |
| 51 | Office for National Statistics. Children's views on well-being and what makes a happy life, UK; 2020. UK; 2020. Available from: https://www.ons.gov.uk/peoplepopulationandcommunity/wellbeing/articles/childrensviewsonwellbeingandwhatmakesahappyifeuk2020/20-10-02. |
| 52 | Public Health Scotland. Children and young people mental health indicators. UK; 2022. Available from: https://publichealthscotland.scot/media/12206/children-and-young-people-mental-health-indicator-set.pdf. |
| 53 | Child Welfare Information Gateway. Measuring and assessing well-being. US; 2020. Available from: https://docs.lib.purdue.edu/cgi/viewcontent.cgi?article=1443&context=jpur. |
| 54 | UNICEF. Understanding child subjective well-being: a call for more data, research and policymaking targeting children. 2021. Available from: https://www.unicef.org/globalinsight/media/2116/file/UNICEF-Global-Insight-Understanding-Child-Subjective-Wellbeing-2021.pdf. |
| 55 | The Children's Society. The good childhood index. UK; 2023. Available from: https://www.childrenssociety.org.uk/information/professionals/good-childhood-index. |
| 56 | Children's services, Government of Alberta. Well-being and resiliency: evaluation framework. Canada: Alberta; 2019. Available from: https://open.alberta.ca/dataset/7d64cad5-f400-4cd3-b7d7-76c3a38ff548/resource/9d3ebd8c-e52f-4964-971e-828d4c3bf21c/download/well-being-resiliency-fmwk-evaluation-march2019.pdf. |
| 57 | World Health Organization. Child Wellbeing Index (WHO-5). WHO Regional Office for Europe: Copenhagen; 1988. Available from: https://www.cocoinitiative.org/sites/default/files/resources/ICI_tools_Wellbeing%20Index%20Guidance.pdf. |
| 58 | Coleman CL, Morrison M, Perkins SK, Brosco JP, Schor EL. Quality of Life and Well-Being for Children and Youth With Special Health Care Needs and their Families: A Vision for the Future. Pediatrics. 2022 Jun 1;149(Suppl 7):e2021056150G. [doi: 10.1542/peds.2021-056150G](https://doi.org/10.1542/peds.2021-056150G). |
| 59 | Turnaround for Children. Well-Being Index - Turnaround for children. US; 2020. Available from: https://turnaroundusa.org/toolbox/mh-campaign-well-being-index/. |
| 60 | Halton Region. Factors that influence a child's social and emotional well-being. Canada: Oakville Ontario; 2023. Available from: https://www.halton.ca/For-Business/Professionals/Early-Learning-and-Child-Care-Professionals/Factors-that-Influence-a-Child-s-Social-and-Emotio. |
| 61 | Sante Publique France. Santé mentale : premiers résultats de l'étude Enabee, chez les enfants de 6 à 11 ans scolarisés du CP au CM2. France; 2023. Available from: https://www.santepubliquefrance.fr/presse/2023/sante-mentale-premiers-resultats-de-l-etude-enabee-chez-les-enfants-de-6-a-11-ans-scolarises-du-cp-au-cm2. |
| 62 | Public Health England. How healthy behaviour supports children's wellbeing. UK: London; 2013. Available from: https://assets.publishing.service.gov.uk/government/uploads/system/uploads/attachment_data/file/232978/Smart_Restart_280813_web.pdf. |
| 63 | Department for Education. State of the nation 2022: children and young people's wellbeing. UK; 2023. Available from: https://assets.publishing.service.gov.uk/government/uploads/system/uploads/attachment_data/file/1134596/State_of_the_nation_2022_-_children_and_young_people_s_wellbeing.pdf. |
| 64 | Educational Psychology Service. Emotional and psychological well-being in children: the standardisation of the stirring children's wellbeing scale. UK; 2015. Available from: https://warwick.ac.uk/fac/sci/med/research/platform/wemwbs/using/faq/scwbs_children_report.pdf. |
| 65 | Nolan A & Smyth E. Risk and protective factors for mental health and wellbeing in childhood and adolescence. Ireland: Dublin; 2021. Available from: https://www.esri.ie/system/files/publications/RS120.pdf. |
| 66 | Children and Young People's Services Committee. CYPSC: Social and emotional well-being: an outline report on social and emotional well-being as it relates to children and young people. Ireland; 2017. Available from: https://www.cypsc.ie/fileupload/Documents/Resourses/Cork/Cork-Sweb-18_04_17.pdf. |
| 67 | UNICEF. Learning through play: strengthening learning through play in early childhood education programmes. US; 2018. Available from: https://ncn.ie/wp-content/uploads/2023/03/UNICEF-Lego-Foundation-Learning-through-Play_2.pdf. |
| 68 | Gregory T, Sincovich A, Brushe M, Finlay-Jones A, Collier LR, Grace B, Sechague Monroy N, Brinkman SA. Basic epidemiology of wellbeing among children and adolescents: A cross-sectional population level study. SSM Popul Health. 2021 Aug 26;15:100907. https://doi.org/10.1016/j.ssmph.2021.100907. |
| 69 | Gregory, T., Brushe, M., Boulton, Z., & Brinkman, S. (2021). Research Snapshot. The Wellbeing of Children and Adolescents in South Australia. Available at https://www.education.sa.gov.au/docs/system-performance/fraser-mustard-centre/wellbeing-of-sa-students-research-snapshot.pdf. |
| 70 | Waddell, C., Shepherd, C.A., & Chen, A. (2013). Creating comprehensive children's mental health indicators for British Columbia. Canadian Journal of Community Mental Health, 32(1), 9-28. https://doi.org/10.7870/cjcmh-2013-003. |
| 71 | Bitsko RH, Claussen AH, Lichstein J, Black LI, Jones SE, Danielson ML, Hoenig JM, Davis Jack SP, Brody DJ, Gyawali S, Maenner MJ, Warner M, Holland KM, Perou R, Crosby AE, Blumberg SJ, Avenevoli S, Kaminski JW, Ghandour RM; Contributor. Mental Health Surveillance Among Children - United States, 2013-2019. MMWR Suppl. 2022 Feb 25;71(2):1-42. https://doi.org/10.15585%2Fmmwr.su7102a1. |
| 72 | Herrman, H., Moodie, R., Saxena, S., University Of Melbourne, Victorian Health Promotion Foundation & World Health Organization. Department Of Mental Health And Substance Abuse. (2005) Promoting Mental Health: Concepts, Emerging Evidence, Practice. [Geneva: World Health Organization, ©] [Pdf] Retrieved from the Library of Congress, https://www.loc.gov/item/2021763240/. |
| 73 | Parkinson J. Establishing a core set of national, sustainable mental health indicators for children and young people in Scotland: Final Report. UK: NHS Health Scotland; 2012. Available from: https://www.healthscotland.scot/media/2249/final-report-on-the-children-and-young-people-s-mental-health-indicators-2012.pdf. |
| 74 | Rodriguez-Ayllon M, Cadenas-Sánchez C, Estévez-López F, Muñoz NE, Mora-Gonzalez J, Migueles JH, Molina-García P, Henriksson H, Mena-Molina A, Martínez-Vizcaíno V, Catena A, Löf M, Erickson KI, Lubans DR, Ortega FB, Esteban-Cornejo I. Role of Physical Activity and Sedentary Behavior in the Mental Health of Preschoolers, Children and Adolescents: A Systematic Review and Meta-Analysis. Sports Med. 2019 Sep;49(9):1383-1410. https://doi.org/10.1007/s40279-019-01099-5. |
| 75 | Kim, E.K., Furlong, M.J., Ng, Z.J., Huebner, E.S. (2020). Child Well-Being and Children's Rights: Balancing Positive and Negative Indicators in Assessments. In: Nastasi, B.K., Hart, S.N., Naser, S.C. (eds) International Handbook on Child Rights and School Psychology. Springer, Cham. https://doi.org/10.1007/978-3-030-37119-7_11. |
| 76 | Halle T & Moore K. Promoting positive and healthy behaviours in children: creating indicators of positive development. US: National Resilience Resource Center; 2012. Available from: https://www.nationalresilienceresource.com/positive_development_2012.pdf. |
| 77 | Blumberg, S.J., Carle, A.C., O'Connor, K.S. et al. Social Competence: Development of an Indicator for Children and Adolescents. Child Ind Res 1, 176–197 (2008). https://doi.org/10.1007/s12187-007-9007-x. |
| 78 | Aber JL, Jones SM. □ Indicators of positive development in early childhood: □ Improving concepts and measures. In Hauser R, Brown B, Prosser W, Stagner M, editors, Indicators of children's well-being. New York: Russell Sage. 1997. p. 395-427. |
| 79 | Furlong, M.J., You, S., Renshaw, T.L. et al. Preliminary Development of the Positive Experiences at School Scale for Elementary School Children. Child Ind Res 6, 753–775 (2013). https://doi.org/10.1007/s12187-013-9193-7. |
| 80 | Ravens-Sieberer U, Auquier P, Erhart M, Gosch A, Rajmil L, Bruil J, Power M, Duer W, Cloetta B, Czemy L, Mazur J, Czimbalamos A, Tountas Y, Hagquist C, Kilroe J; European KIDSCREEN Group. The KIDSCREEN-27 quality of life measure for children and adolescents: psychometric results from a cross-cultural survey in 13 European countries. Qual Life Res. 2007 Oct;16(8):1347-56. https://doi.org/10.1007/s11136-007-9240-2. |
| 81 | Minkkinen, J. The Structural Model of Child Well-Being. Child Ind Res 6, 547–558 (2013). https://doi.org/10.1007/s12187-013-9178-6. |
| 82 | Newland, L.A., Lawler, M.J., Giger, J.T. et al. Predictors of Children's Subjective Well-Being in Rural Communities of the United States. Child Ind Res 8, 177–198 (2015). https://doi.org/10.1007/s12187-014-9287-x. |
| 83 | Bernard, M.E., Stephanou, A. Ecological Levels of Social and Emotional Wellbeing of Young People. Child Ind Res 11, 661–679 (2018). https://doi.org/10.1007/s12187-017-9466-7. |
| 84 | Schuksmith J, Spratt J, Philip K, McNaughton R. A critical review of the literature on children and young people's views of the factors that influence their mental health. UK: Edinburgh NHS Health Scotland; 2009. Available from: https://www.healthscotland.scot/media/2247/a-critical-review-of-the-literature-on-children-and-young-people-s-views-of-the-factors-that-influence-their-mental-health-2009.pdf. |
| 85 | O'Connor, M., Cloney, D., Kvalsvig, A., & Goldfeld, S. (2019). Positive mental health and academic achievement in elementary school: New evidence from a matching analysis. Educational Researcher, 48(4), 205–216. https://doi.org/10.3102/0013189X19848724. |
| 86 | de Lannoy, L., Barbeau, K., Vanderloo, L. M., Goldfield, G., Lang, J. J., MacLeod, O., & Tremblay, M. S. (2023). Evidence supporting a combined movement behavior approach for children and youth's mental health—A scoping review and environmental scan. *Mental Health and Physical Activity*, 24, 1–15. https://doi.org/10.1016/j.mhpa.2023.100511. |
| 87 | Oberle E, Guhn M, Gadermann AM, Thomson K, Schonert-Reichl KA. Positive mental health and supportive school environments: A population-level longitudinal study of dispositional optimism and school relationships in early adolescence. *Soc Sci Med*. 2018 Oct;214:154-161. https://doi.org/10.1016/j.socscimed.2018.06.041. |
| 88 | Maughan B, Collishaw S, Meltzer H, Goodman R. Recent trends in UK child and adolescent mental health. *Soc Psychiatry Psychiatr Epidemiol*. 2008 Apr;43(4):305-10. https://doi.org/10.1007/s00127-008-0310-8. |
| 89 | Smith NDW, Suldo SM, Hearon BV, Ferron J. An Application of the Dual-Factor Model of Mental Health in Elementary School Children: Examining Academic Engagement and Social Outcomes. *Journal of Positive Psychology & Wellbeing*. 2020;4(1):49-68. https://journalppw.com/index.php/jppw/article/view/61 |
| 90 | de Carvalho, J.S., Pereira, N.S., Pinto, A.M. et al. Psychometric Properties of the Mental Health Continuum-Short Form: A Study of Portuguese Speaking Children/Youths. *J Child Fam Stud* 25, 2141–2154 (2016). https://doi.org/10.1007/s10826-016-0396-7. |
| 91 | Ottova, V., Hjern, A., Rasche, C., & Ravens-Sieberer, U. (2012). Child mental health measurement : reflections and future directions. https://doi.org/10.5772/37830. |
| 92 | Prilleltensky, I., Nelson, G., & Peirson, L. (2001). The role of power and control in children's lives: An ecological analysis of pathways toward wellness, resilience and problems. *Journal of Community & Applied Social Psychology*, 11(2), 143–158. https://doi.org/10.1002/casp.616. |
| 93 | Jones DE, Greenberg M, Crowley M. Early Social-Emotional Functioning and Public Health: The Relationship Between Kindergarten Social Competence and Future Wellness. *Am J Public Health*. 2015 Nov;105(11):2283-90. https://doi.org/10.2105/ajph.2015.302630. |
| 94 | Dudovitz RN, Thomas K, Shah MD, Szilagyi PG, Vizueta N, Vangala S, Shetgiri R, Kapteyn A. School-Age Children's Wellbeing and School-Related Needs During the COVID-19 Pandemic. *Acad Pediatr*. 2022 Nov-Dec;22(8):1368-1374. https://doi.org/10.1016/j.acap.2022.01.015. |
| 95 | Julia Buzaud, Zoé Perron, Kevin Diter, Claude Martin. Les mesures du bien-être des enfants Revue de littérature sur les grandes enquêtes internationales. *Revue des politiques sociales et familiales*, 2019. https://dx.doi.org/10.3406/caf.2019.3350. |
| 96 | Office for National Statistics. "Children's well-being and social relationships, UK: 2018. UK; 2018. Available from: https://backup.ons.gov.uk/wp-content/uploads/sites/3/2018/03/Children-s-well-being-and-social-relationships-UK-2018-1.pdf. |
| 97 | City of Westminster. Westminster Health and Wellbeing Board. UK; 2017. Available from: https://committees.westminster.gov.uk/documents/s23716/6%20-%20Draft%20Annual%20Director%20of%20Public%20Health%20Report.pdf. |
| 98 | Barry, M., & Friedli, L. (2010). The influence of social, demographic and physical factors on positive mental health in children, adults and older people. In C. L. Cooper, J. Field, U. Goswami, R. Jenkins, & B. J. Sahakian (Eds.), *Mental capital and wellbeing* (pp. 475–484). Wiley Blackwell. |
| 99 | Dobia, B., & O'Rourke, V. (2011). Promoting the Mental Health and Wellbeing of Indigenous Children in Australian Primary Schools. Canberra, A.C.T.: Commonwealth of Australia. Available from: https://researchdirect.westernsydney.edu.au/islandora/object/uws:16589/. |
| 100 | Stewart, D., Sun, J., Patterson, C., Lemerle, K., & Hardie, M. (2004). Promoting and Building Resilience in Primary School Communities: Evidence from a Comprehensive ‘Health Promoting School’ Approach. *International Journal of Mental Health Promotion*, 6(3), 26–33. https://doi.org/10.1080/14623730.2004.9721936 . |
| 101 | Canadian Public Health Association. Canada; CPHA: 2019. What's in the best interest of the child? A public health perspective on access to unstructured play for children in Canada. Available from: https://www.cpha.ca/sites/default/files/uploads/resources/play/playproject-wickedproblems-e.pdf. |
| 102 | Canadian Institute for Health Information. Child and Youth Health and Well-Being Indicators Project: CIHI and B.C. PHO Joint Summary Report, February 2013. Ottawa, ON: CIHI; 2013. Available from: https://www2.gov.bc.ca/assets/gov/health/about-bc-s-health-care-system/office-of-the-provincial-health-officer/reports-publications/special-reports/child-health-2013.pdf. |
| 103 | OECD (2021), *Measuring What Matters for Child Well-being and Policies*, OECD Publishing, Paris. https://doi.org/10.1787/e82fded1-en. |
| 104 | Morrison W & Peterson P. Pan-Canadian Joint Consortium for School Health: Positive Mental Health Toolkit. Canada; JSCH: 2017. Available from: http://www.jcsh-cces.ca/images/upload/JCSHPositiveMentalHealthToolkit.pdf. |

### Study characteristics

An overview of the characteristics for the sources included in the scoping review can be found in Table 3. Many of the sources were from the US (n=28) and UK (n=21), with 18 that were international (included at least one Western country). Other countries included Australia (n=13), Canada (n=9), Ireland (n=3), Brazil (n=2), and Spain (n=2). There was one source from each of the following countries: France, Germany, Hungary, Italy, Netherlands, Portugal, Sweden, and Switzerland. The most common types of evidence included were government reports (n=26) and cross-sectional studies (n=24). This was followed by evidence from non-governmental organizations (e.g., World Health Organization [WHO], OECD; n=10), reviews (n=10), book chapters (n=7), validation studies (n=7), research reports (n=5), qualitative studies (n=4), cohort studies (n=3), conceptual/theoretical papers (n=3), longitudinal studies (n=3), and position papers (n=2). The majority of the evidence was published between 2010 and 2019 (52.0%), followed by 2020-onwards (32.7%). The majority of the peer-reviewed studies only covered the middle childhood stage (ages 6-11; 64.1%), whereas the majority of the grey literature sources touched on the overall stage of childhood (including both early and middle childhood; 56.9%). Few sources examined early childhood only (age 5 and under; n=5). Lastly, only one source was published in French and the rest (n=103) were published in English.

**Table 3.**
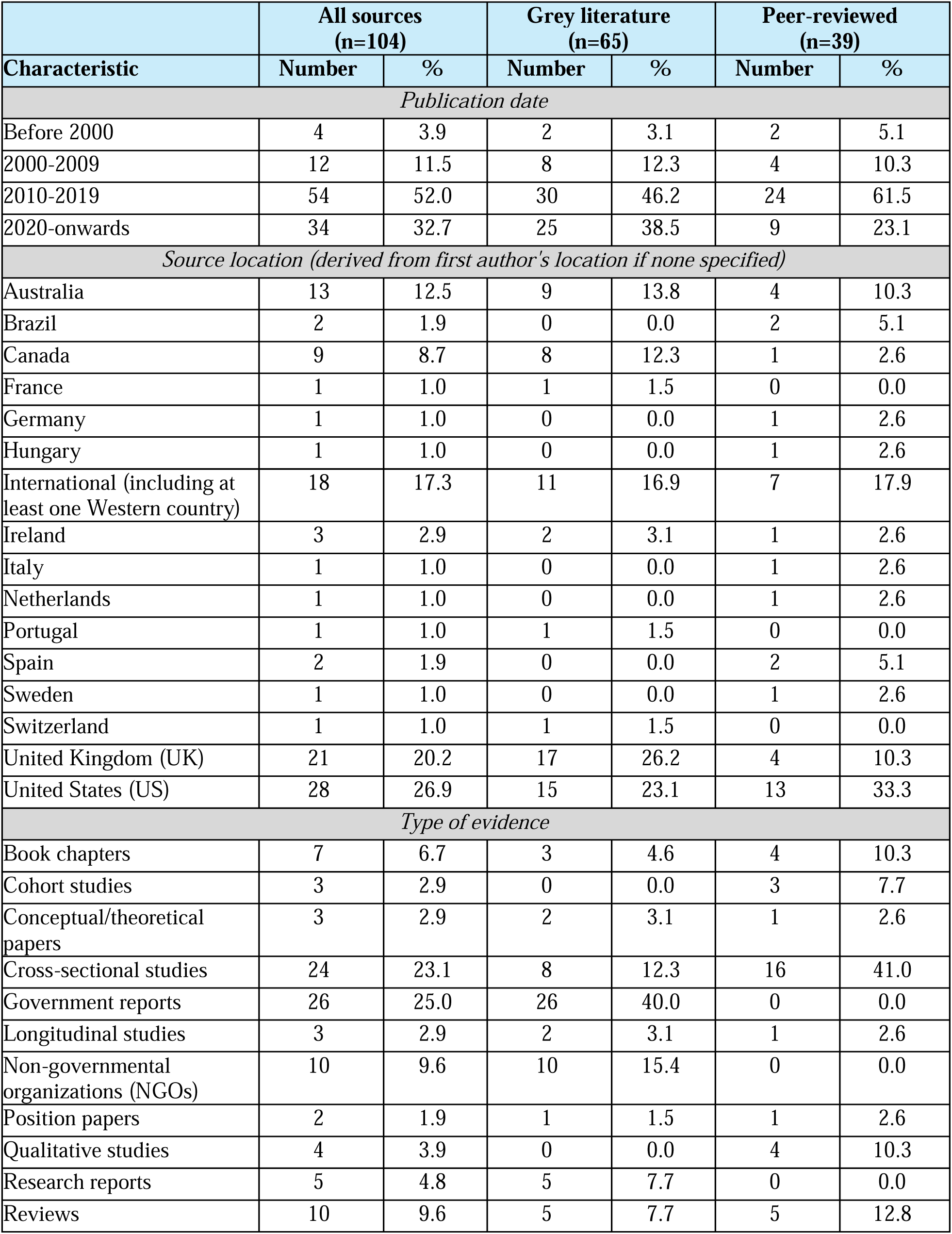
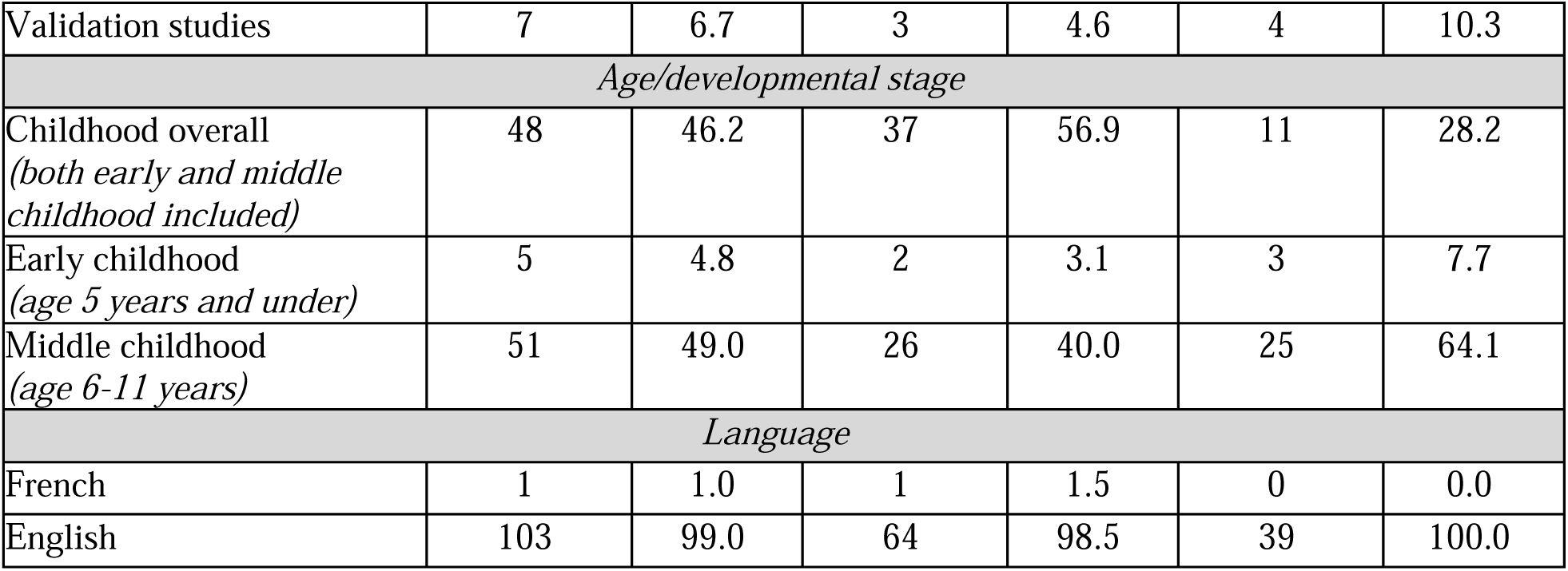
Characteristics of sources included in scoping review.

### Theories mentioned in the introduction/background of sources

The majority (74.0%, n=77) of sources mentioned at least one theory in the introduction/background. Of those sources, 62.3% (n=48) used more than one theory to conceptualize child PMH. The most commonly identified theories for children were positive psychology theories (n=31), the dual continuum of mental health (n=23), child rights theories (n=17), developmental theories (n=16), the multidimensional approach to subjective well-being (n=15) and the socio-ecological theory/model (n=14). Twenty-seven articles did not provide any theoretical rationale in their introduction/background. A list of all theories (including a brief explanation for each), associated source IDs for each source that mentioned the theory, and the total number of sources per theory can be found in Table 4.

**Table 4.**
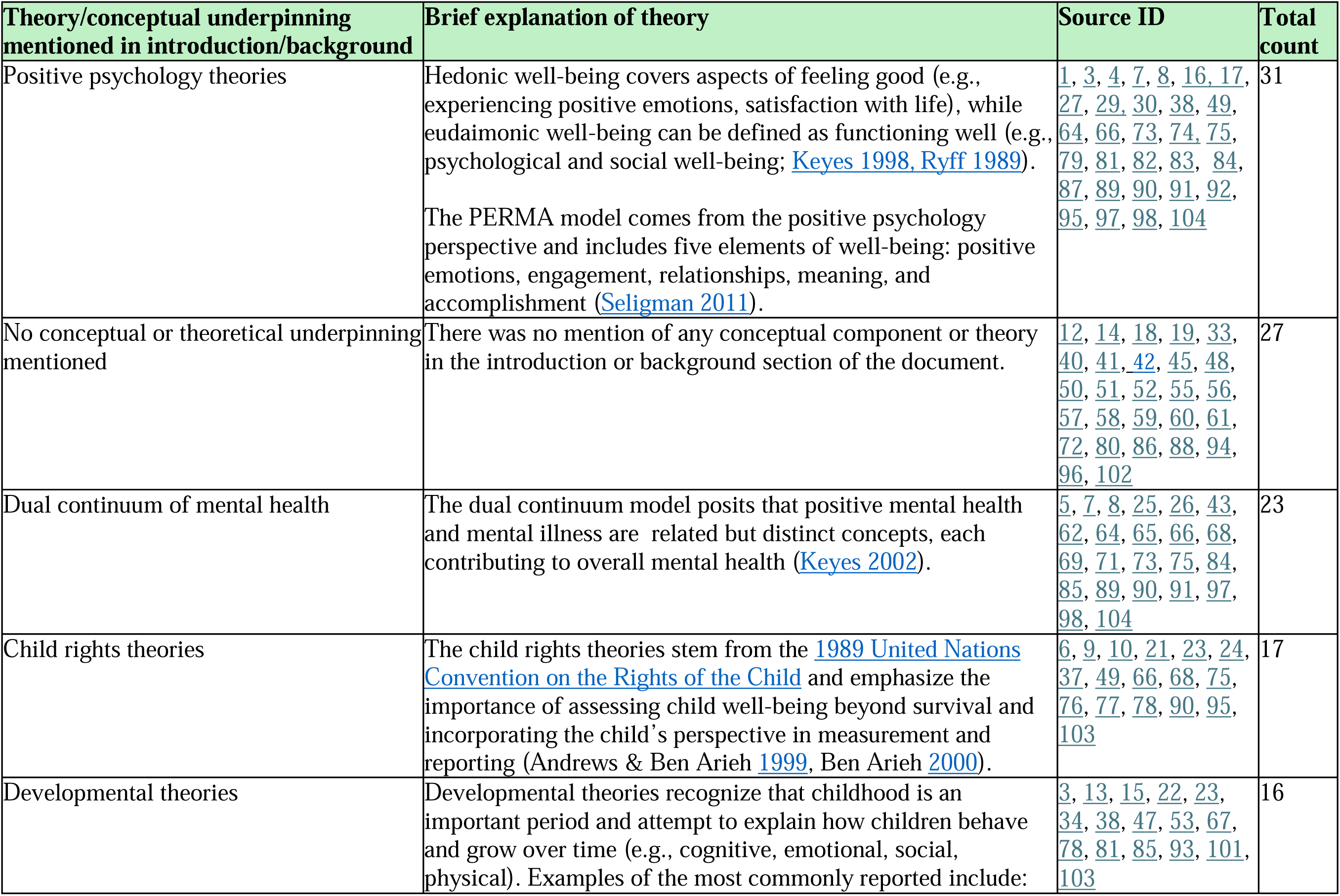
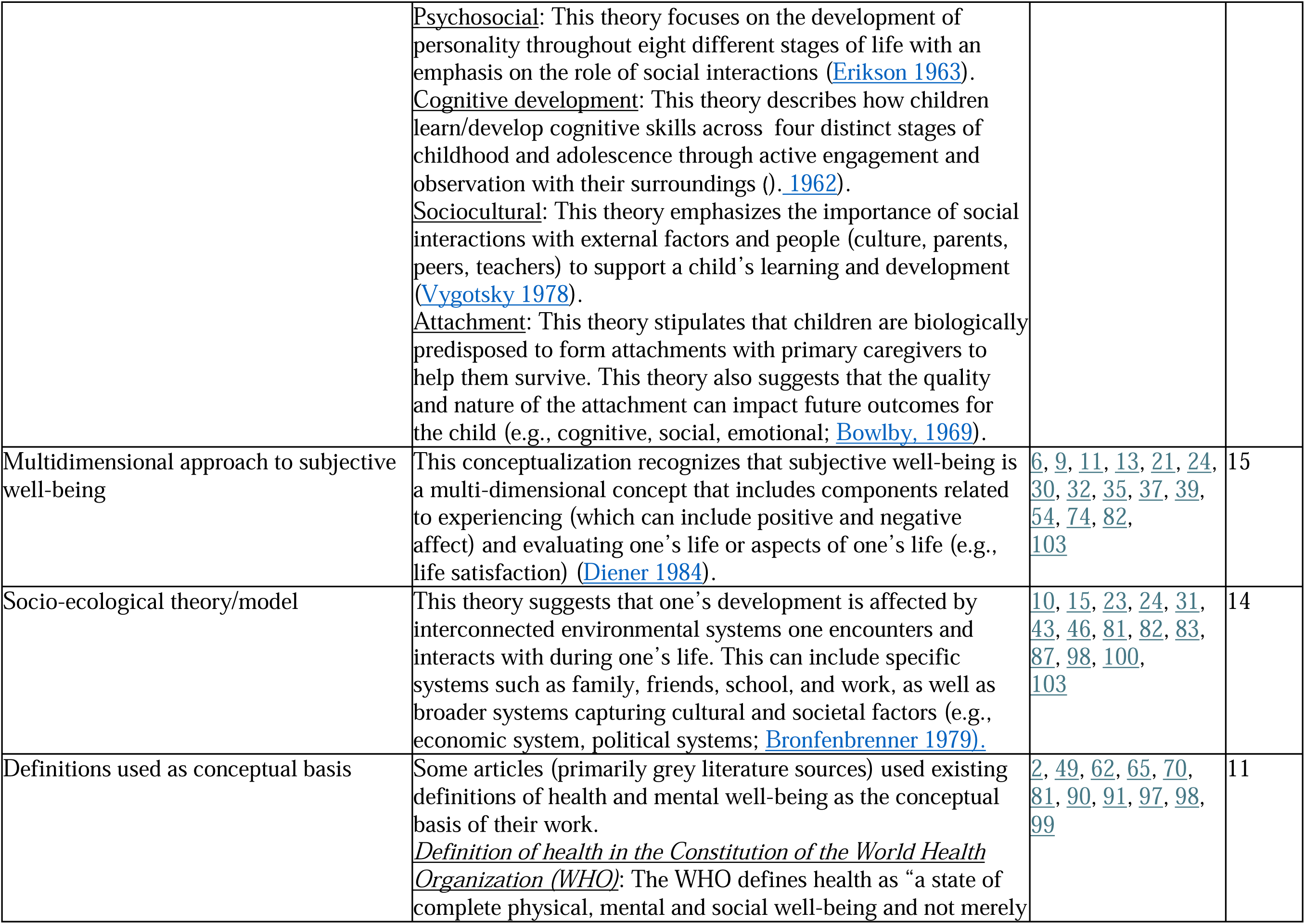
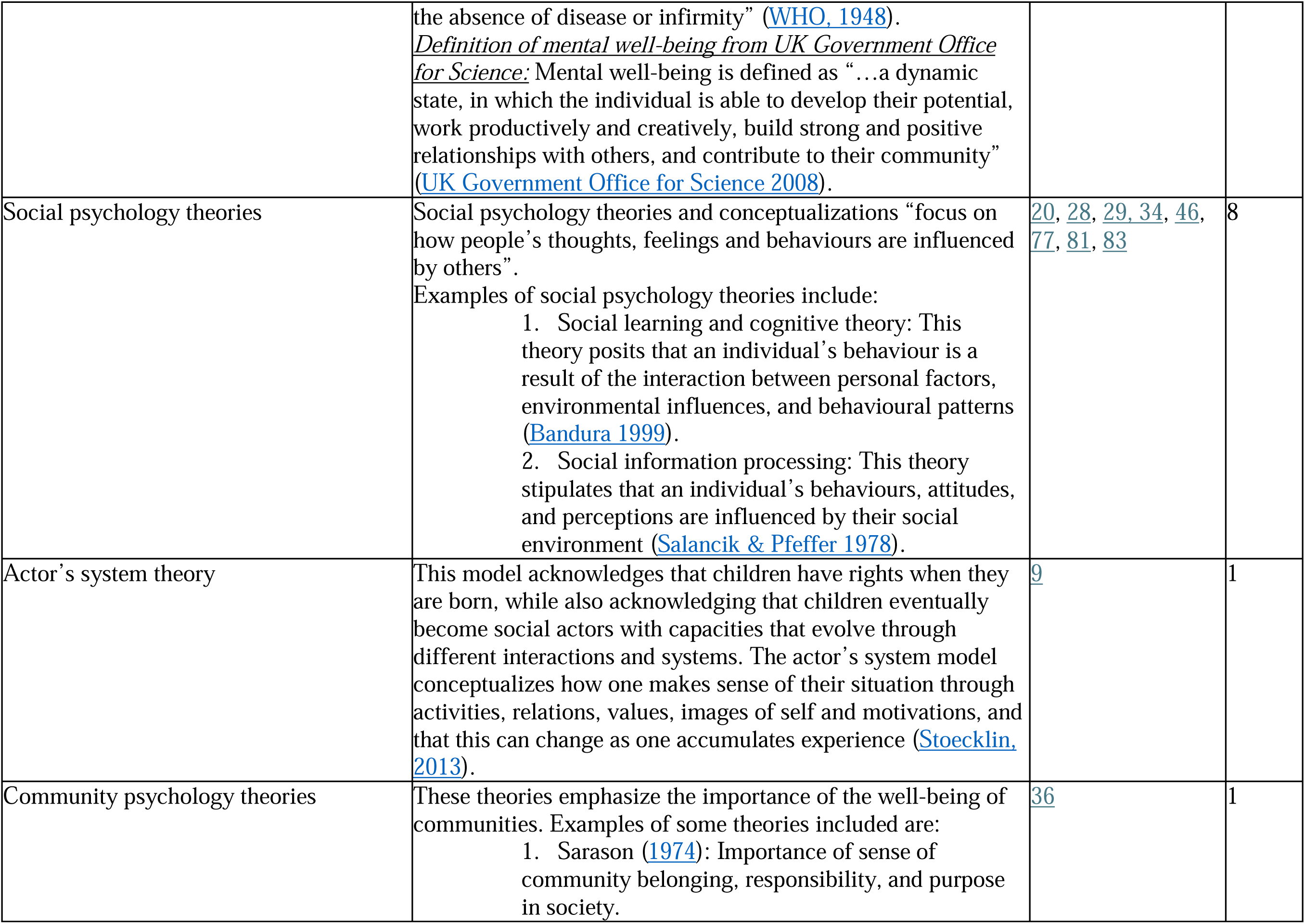
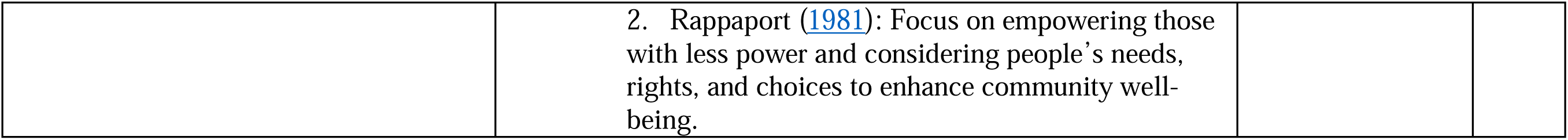
Theories mentioned in introduction/background, brief explanation of theory, and total count.

### PMH concepts/outcomes and instruments

The included sources covered different PMH concepts and outcomes, which were grouped into four different categories. Close to three quarters of sources (74.0%, n=77) examined hedonic well-being (including subjective/emotional/affective well-being). Other PMH concepts/outcomes that emerged through thematic analysis were psychological well-being (n=41), social well-being (n=30), and social emotional learning and/or positive development (n=19). A brief explanation of each PMH concept/outcome, associated source IDs for each source that mentioned the PMH concept/outcome, and the total number of sources per category can be found in Table 5.

**Table 5.**
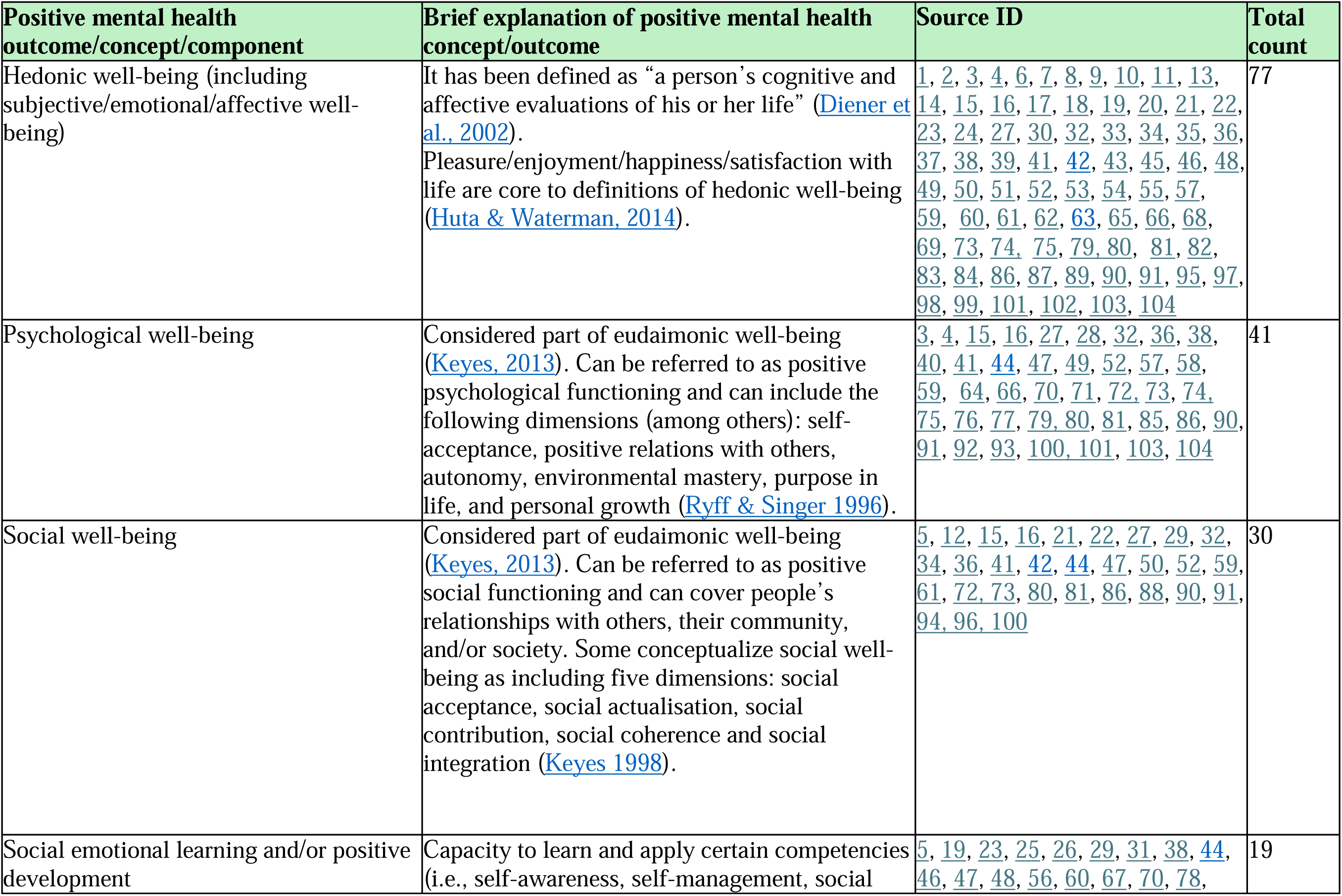
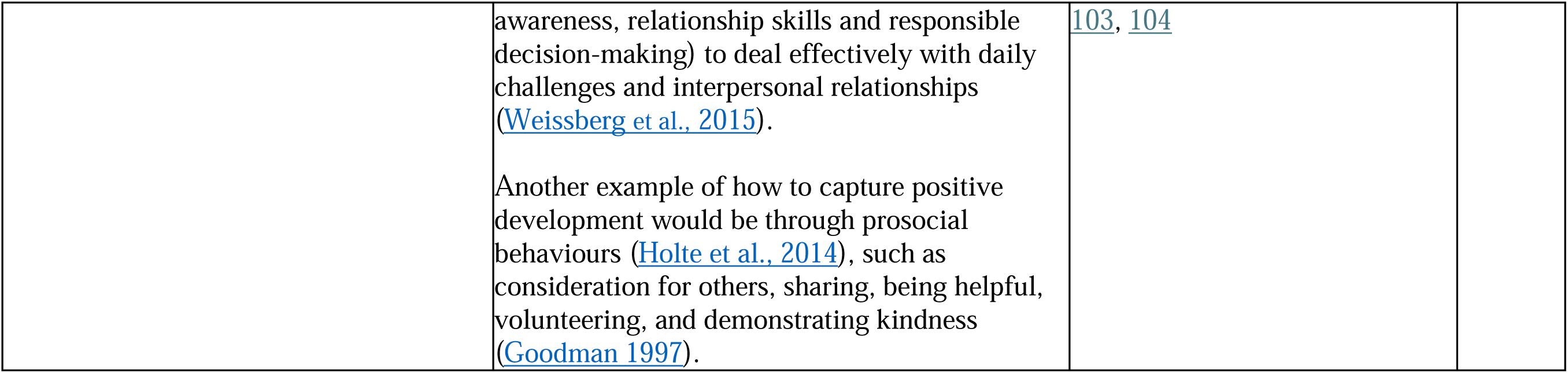
Positive mental health outcome/concept/component extracted, brief explanation for each and total count.

| Positive mental health outcome/concept/component | Brief explanation of positive mental health concept/outcome | Source ID | Total count |
| --- | --- | --- | --- |
| Hedonic well-being (including subjective/emotional/affective well-being) | It has been defined as “a person’s cognitive and affective evaluations of his or her life” (Diener et al., 2002).<br>Pleasure/enjoyment/happiness/satisfaction with life are core to definitions of hedonic well-being (Huta & Waterman, 2014). | 1, 2, 3, 4, 6, 7, 8, 9, 10, 11, 13, 14, 15, 16, 17, 18, 19, 20, 21, 22, 23, 24, 27, 30, 32, 33, 34, 35, 36, 37, 38, 39, 41, 42, 43, 45, 46, 48, 49, 50, 51, 52, 53, 54, 55, 57, 59, 60, 61, 62, 63, 65, 66, 68, 69, 73, 74, 75, 79, 80, 81, 82, 83, 84, 86, 87, 89, 90, 91, 95, 97, 98, 99, 101, 102, 103, 104 | 77 |
| Psychological well-being | Considered part of eudaimonic well-being (Keyes, 2013). Can be referred to as positive psychological functioning and can include the following dimensions (among others): self-acceptance, positive relations with others, autonomy, environmental mastery, purpose in life, and personal growth (Ryff & Singer 1996). | 3, 4, 15, 16, 27, 28, 32, 36, 38, 40, 41, 44, 47, 49, 52, 57, 58, 59, 64, 66, 70, 71, 72, 73, 74, 75, 76, 77, 79, 80, 81, 85, 86, 90, 91, 92, 93, 100, 101, 103, 104 | 41 |
| Social well-being | Considered part of eudaimonic well-being (Keyes, 2013). Can be referred to as positive social functioning and can cover people’s relationships with others, their community, and/or society. Some conceptualize social well-being as including five dimensions: social acceptance, social actualisation, social contribution, social coherence and social integration (Keyes 1998). | 5, 12, 15, 16, 21, 22, 27, 29, 32, 34, 36, 41, 42, 44, 47, 50, 52, 59, 61, 72, 73, 80, 81, 86, 88, 90, 91, 94, 96, 100 | 30 |
| Social emotional learning and/or positive development | Capacity to learn and apply certain competencies (i.e., self-awareness, self-management, social | 5, 19, 23, 25, 26, 29, 31, 38, 44, 46, 47, 48, 56, 60, 67, 70, 78, | 19 |
|  | awareness, relationship skills and responsible decision-making) to deal effectively with daily challenges and interpersonal relationships (Weissberg et al., 2015).<br>Another example of how to capture positive development would be through prosocial behaviours (Holte et al., 2014), such as consideration for others, sharing, being helpful, volunteering, and demonstrating kindness (Goodman 1997). | 103, 104 |  |

There are numerous instruments available in the literature that can be used to report on aspects of PMH among children. Table 6 includes measures capturing aspects of PMH that were extracted from the sources and categorized as self-rated (i.e., child-rated) or other-rated (e.g., parent, teacher).

**Table 6.** Instruments capturing positive mental health outcomes/concepts that were extracted for children 11 years and under.

| Self-rated | Other-rated |
| --- | --- |
| Brief Multidimensional Student's Life Satisfaction Scale (ages 8-11) | Early year Development Instrument (EDI) (teacher-rated for children aged 3.5-6.5) |
| Cantril's ladder (life satisfaction) (ages 11-15) | Happiness scales (e.g., average parent<sup>a</sup>-reported happiness of children aged 5-11) |
| Child Health Questionnaire (CHQ) (ages 5-18) | KIDSCREEN (10-item, 27-item, and 52 –item versions) (ages 8-18) |
| Children's World's Subjective Well-being scale (CW-SWBS) (ages 7-10) | Parent-rated child behaviour (e.g., child usually or always smiles and laughs) |
| Children's Worlds 10-year old Survey - adapted for 5th graders | Prosocial Subscale of Strengths and Difficulties Questionnaire (SDQ; parent-and teacher-rated for children/youth aged 2-17) |
| Child and Youth Resilience Measure (CYRM) - Child version (ages 5-9) | Social Competence Scale (parent-reported for children/youth aged 6-17, included in National Survey of Children's Health [NSCH]) |
| Good Childhood Index (ages 10-17) | Social and Emotional Well-Being Survey (teacher-rated for students aged 3-18) |
| Happiness (e.g., frequency, happiness with aspects of life) (ages 6-17) | The Australian Early Development Census (AEDC) (teacher-rated for children aged 5-6) |
| How I Feel About My School (HIFAMS) questionnaire (ages 4-12) |  |
| Huebner's Life Satisfaction Scale (ages 8-18) |  |
| KIDSCREEN (10-item, 27-item, and 52 –item versions) (ages 8-18) |  |
| Mental Health Continuum Short Form (adapted for children ages 7+) |  |
| Middle years Development Instrument (MDI) (grades 4-8) |  |
| Pediatric Quality of Life (PedsQL) Generic Core Scale (ages 5-18) |  |
| Personal Wellbeing Index–School Children (PWI-SC) (age 11+) |  |
| Prosocial Subscale of Strengths and Difficulties Questionnaire (SDQ) (ages 11-17) |  |
| Social and Emotional Well-Being Survey (student) (ages 7+) |  |
| Stirling Children's Wellbeing Scale (SCWBS) (ages 8-15) |  |
| Student Well-Being Index (ages 8+) |  |
| The World Health Organization – Five Well-Being Index (WHO-5) (ages 9+) |  |

### PMH risk and protective factors

Many risk and protective factors were extracted from the included studies. These included factors at the individual level (e.g., healthy living behaviours like physical activity and sleep), family level (e.g., family dynamics), community level (e.g., school environment/experiences), and society level (e.g., access to services). The full list of high-level determinants categorized across the four domains with examples can be found in Table 7.

**Table 7.**
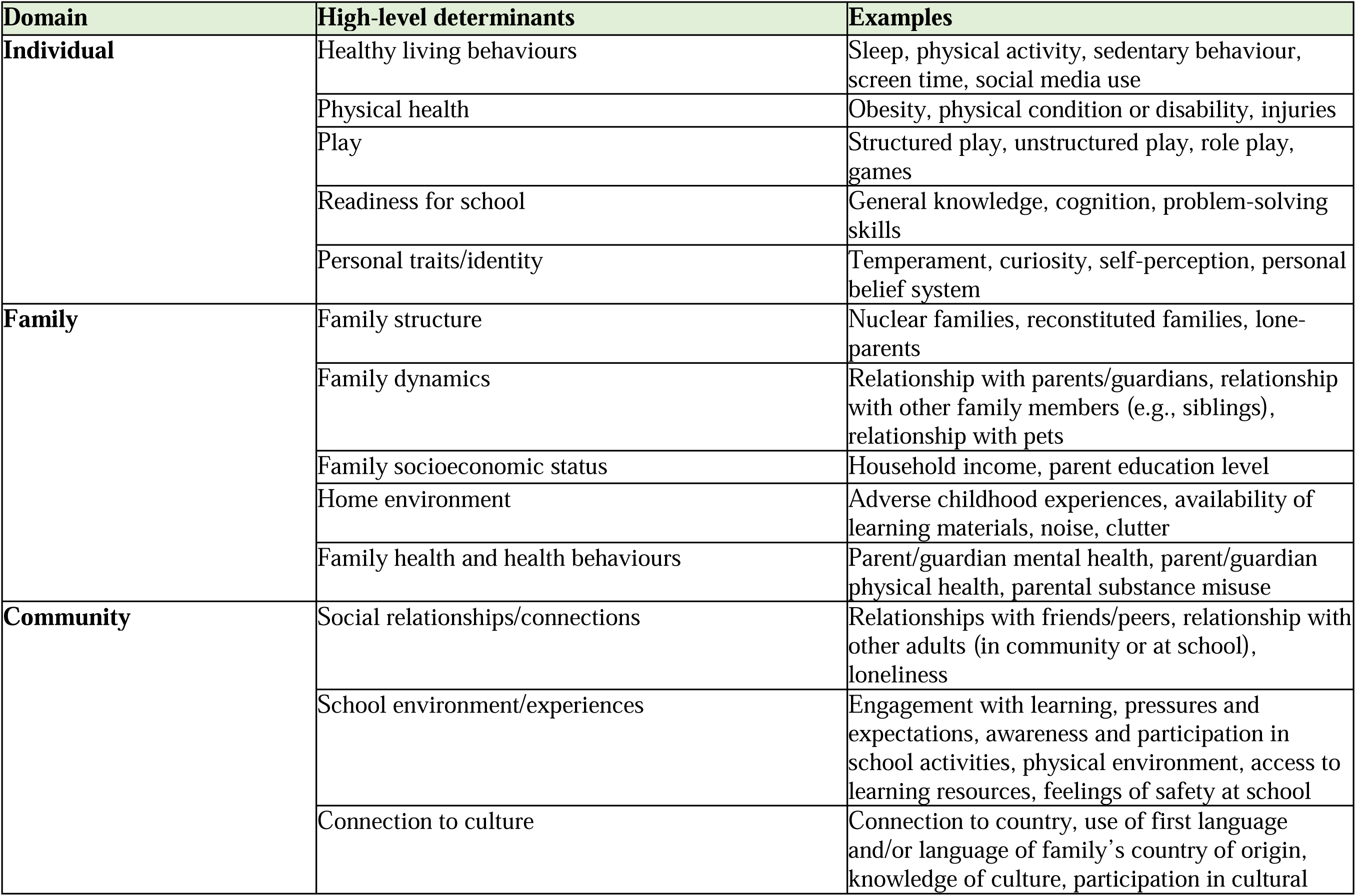
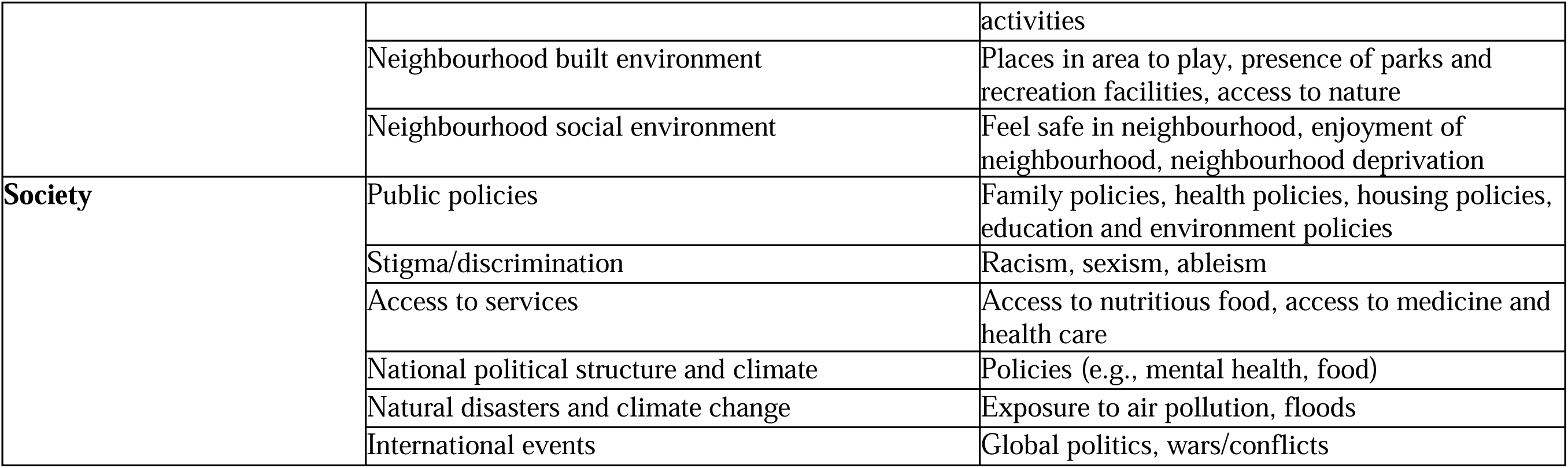
High-level determinants categorized under four socio-ecological domains.

| Domain | High-level determinants | Examples |
| --- | --- | --- |
| **Individual** | Healthy living behaviours | Sleep, physical activity, sedentary behaviour, screen time, social media use |
|  | Physical health | Obesity, physical condition or disability, injuries |
|  | Play | Structured play, unstructured play, role play, games |
|  | Readiness for school | General knowledge, cognition, problem-solving skills |
|  | Personal traits/identity | Temperament, curiosity, self-perception, personal belief system |
| **Family** | Family structure | Nuclear families, reconstituted families, lone-parents |
|  | Family dynamics | Relationship with parents/guardians, relationship with other family members (e.g., siblings), relationship with pets |
|  | Family socioeconomic status | Household income, parent education level |
|  | Home environment | Adverse childhood experiences, availability of learning materials, noise, clutter |
|  | Family health and health behaviours | Parent/guardian mental health, parent/guardian physical health, parental substance misuse |
| **Community** | Social relationships/connections | Relationships with friends/peers, relationship with other adults (in community or at school), loneliness |
|  | School environment/experiences | Engagement with learning, pressures and expectations, awareness and participation in school activities, physical environment, access to learning resources, feelings of safety at school |
|  | Connection to culture | Connection to country, use of first language and/or language of family's country of origin, knowledge of culture, participation in cultural |
|  |  | activities |
|  | Neighbourhood built environment | Places in area to play, presence of parks and recreation facilities, access to nature |
|  | Neighbourhood social environment | Feel safe in neighbourhood, enjoyment of neighbourhood, neighbourhood deprivation |
| **Society** | Public policies | Family policies, health policies, housing policies, education and environment policies |
|  | Stigma/discrimination | Racism, sexism, ableism |
|  | Access to services | Access to nutritious food, access to medicine and health care |
|  | National political structure and climate | Policies (e.g., mental health, food) |
|  | Natural disasters and climate change | Exposure to air pollution, floods |
|  | International events | Global politics, wars/conflicts |

## Discussion

This scoping review identified relevant theories, concepts/outcomes, and determinants of PMH in children 11 years and under that can be used to extend public health surveillance of PMH in Western countries.

### Theories

The majority of the sources on child PMH had theoretical/conceptual underpinnings in their introduction/background sections. Many of the commonly reported theories align with the Public Health Agency of Canada’s conceptualization and routine reporting of PMH (Orpana et al., 2016). Indeed, Canada’s current PMHSIF (for youth and adults) follows the dual continuum of mental health (i.e., differentiates PMH from mental illness), is based on positive psychology theories (e.g., includes hedonic and eudaimonic well-being outcomes), takes a multidimensional approach to measuring and reporting subjective well-being (i.e., includes separate indicators for happiness and life satisfaction), and incorporates a socio-ecological model in its reporting (i.e., acknowledges that one’s PMH can be impacted by external systems by also monitoring family, community, and society determinants). These results support the applicability of much of Canada’s current conceptual framework of PMH (Orpana et al., 2016) to childhood.

This scoping review also uncovered theories that are particularly relevant to childhood. The child rights theories stem from the 1989 United Nations Convention on the Rights of the Child and emphasize the importance of assessing child well-being beyond survival, which can be accomplished by reporting on a child’s PMH, and incorporating the child’s perspective in measurement and reporting (Andrews & Ben-Arieh, 1999; Ben-Arieh, 2000; WHO, 1998). Developmental theories recognize that childhood is a period with unique needs and challenges at different stages (e.g., early childhood vs. middle childhood) that should be assessed accordingly and can impact future well-being outcomes (Bowlby J, 1969; Erikson E, 1963; Piaget J, 1962; Vygotsky LS, 1978). With regards to incorporating child-relevant theories in a Canadian child PMHSIF, two key elements will be critical: 1) incorporating children’s perspective in the measurement and reporting of PMH outcomes (i.e., ensuring that self-reported PMH measures are included) and 2) only including instruments that have been validated among children to ensure that they are age-/stage-sensitive.

It is worth noting that most of the sources used more than one theory to conceptualize child PMH. Furthermore, given the variety of types of peer-reviewed and grey literature sources that were included in the scoping review, the theoretical/conceptual information included in each source varied greatly. For example, some types of evidence had less detailed information on theories and tended to refer to a certain definition as the foundation rather than a specific theory, while others identified specific sub-theories (e.g., attachment theory, psychosocial).

Although there were differences in the theoretical/conceptual information, many of the theories were analogous. For example, the frequent emphasis on the importance of social interactions in the developmental (Bowlby J, 1969; Erikson E, 1963; Piaget J, 1962; Vygotsky LS, 1978), social psychology (Bandura A, 1999; Salancik & Pfeffer, 1978), actor’s system (Stoecklin D, 2013), and community psychology theories (Rappaport J, 1981; Sarason SB, 1974) are akin to Bronfenbrenner’s ecological theory (Bronfenbrenner U, 1979) that also emphasizes the influence of social encounters with inter-connected systems (e.g., family, friends, school, work, economic, political) on human development. Another example is the positive psychology theories (Keyes CLM, 1998; Ryff CD, 1989; Seligman MEP, 2011) and the multidimensional approach to subjective well-being (Diener E, 1984) as hedonic well-being is often construed as including life satisfaction and positive affect (Huta & Waterman, 2014). There is also overlap in some of the definitions that were extracted from the literature with some of the theories. For instance, the WHO definition of health (WHO, 1948) makes a distinction between illness and well-being like the dual continuum model of mental health (Keyes CL, 2002).

Overall, our synthesis suggests that there does not appear to be one overarching ‘gold standard’ theory for conceptualizing child PMH. Rather, it seems like incorporating a mix of different theories from different disciplines may provide the most comprehensive and robust conceptual representation of child PMH. The six principles underlying the OECD conceptual framework for child social emotional well-being provide an example of an interdisciplinary theoretical approach with relevance to identified theories: 1) multidimensional (e.g., *dual continuum of mental health, positive psychology theories, multidimensional approach to subjective well-being*), 2) reflect children’s lives today and tomorrow (e.g., *developmental theories, social psychology theories*), 3) be age- or stage-sensitive (e.g., *developmental theories*), 4) integrate children’s views and perspectives (e.g., *child rights theories*), 5) capture children’s environments (families, schools, neighbourhoods, communities; e.g., *socio-ecological theory/model*), and 6) include child-related public policies (e.g., *socio-ecological theory/model*) (OECD, 2021). The current PMHSIF for youth and adults in Canada already integrates most of the commonly mentioned theories and the addition of the child-relevant components (i.e., be age- or stage-sensitive, and integrate children’s views and perspectives) that reflect the developmental and child rights theories would require minimal changes. These adaptations to the PMHSIF would ensure it embodies both the theoretical/conceptual evidence gathered from this scoping review and would align with OECD conceptual framework for child social emotional well-being.

### PMH concepts and outcomes

According to the evidence gathered, PMH among children is composed of different components including hedonic (subjective/emotional/affective) well-being, eudaimonic (psychological and social) well-being, and social emotional learning and/or positive development. Hedonic and eudaimonic well-being are not concepts unique to children; they also extend to youth and adults, as seen in Canada’s PMHSIF (Orpana et al., 2016; PHAC, 2024). Importantly, hedonic and eudaimonic well-being outcomes are aligned with key conceptual theories that were highlighted in the section above: positive psychology theories (Keyes CLM, 1998; Ryff CD, 1989; Seligman MEP, 2011), dual continuum of mental health (Keyes CL, 2002), multidimensional approach to subjective well-being (Diener E, 1984) and social psychology theories (Bandura A, 1999; Salancik & Pfeffer, 1978). The concept of positive development/social emotional learning emerged through the extraction and synthesis of results. While this is not currently captured in the PMHSIF (PHAC, 2024), the definition for this concept (Weissberg RP et al., 2015) is comparable to the definitions of eudaimonic well-being (i.e., functioning well) (Keyes CLM, 1998; Ryff & Singer, 1996). This potential overlap in concepts could simply be due to differences in terminology. The terminology used to refer to PMH is not standardized (Vaingankar JA et al., 2022), which can cause discrepancies in the terms used to refer to similar constructs in the literature, silo information based on different disciplines/researchers’ preferred terms, and conflate findings involving different constructs. Nevertheless, the majority of sources reported on concepts of hedonic (n=77) and/or eudaimonic (n=53) well-being for child PMH, which directly aligns with the concepts currently included in Canada’s PMHSIF (PHAC, 2024).

### PMH instruments

Numerous instruments used to measure various PMH concepts and outcomes were identified through this scoping review. While the evidence demonstrated that the PMH concepts and outcomes for youth and adults are also applicable to children, there are differences in how they are operationalized. For example, the prosocial subscale of the Strengths and Difficulties Questionnaire (Goodman R, 1997) can be used to measure positive development for children, whereas this scale is not applicable to adults. The need to report on concepts using age-appropriate measures/methodologies has been highlighted in a recent OECD report (Soffia & Turner, 2021). Another key distinction in the measurement of PMH concepts and outcomes for children is that, in addition to self-rated instruments, other-rated instruments (e.g., parent, person most knowledgeable, teacher) are commonly used. There appears to be general agreement in the literature that using a multi-informant approach (i.e., having a mix other-rated and self-rated assessments) provides the most comprehensive picture of child well-being. The majority of the instruments applied to middle childhood (ages 6-11), with fewer available for early childhood (ages 5 and under). Additionally, some of these instruments are currently being used to report on children’s PMH internationally (Children’s Worlds; Ravens-Sieberer et al., 2007) and in countries such as Portugal (Carvalho JS et al., 2016), Australia (Howells et al., 2024), the US (Blumberg et al., 2008; CDC, 2024; Huebner ES, 1994; Moore KA et al., 2012; Ravens-Sieberer et al., 2014), and the UK (Allen et al., 2018; Goodman R, 2001; Hope et al., 2019; The Children’s Society, 2010), with even some in Canada (Janus & Offord, 2007).

Since this scoping review only categorized the instruments based on one criteria (i.e., informant), further work is needed to assess them more thoroughly. For instance, evaluating which PMH components the instruments are capturing could be informative. Moreover, certain methodological concerns, such as the validation of specific instruments among children (e.g., the Personal Wellbeing Index – School Children for those under 10 years old) (Bellamy L, 2020) and cross-national comparability of certain measures (Casas & Rees, 2015), have been raised. Validation studies may be needed to confirm the reliability and validity of some instruments among Canadian children.

While a more in-depth review of measures is needed, we are aware of three potential measures of child PMH that are currently available in national population-based health surveys in Canada. Specifically, there is an other-rated measure of child mental health (*available in the Canadian Health Survey for Children and Youth*), an other-rated measure of the prosocial subscale of the Strengths and Difficulties Questionnaire (*available in the Canadian Health Measures Survey*), and a self-reported measure (for children aged 6-11 years) of happiness (*available in the Canadian Health Measures Survey*) (Hoffmann et al., 2020; Statistics Canada, 2023; Statistics Canada, 2024; Turner et al., 2023). There remains insufficient data on other PMH components (such as psychological well-being), creating a gap in Canadian evidence. Given that the regular national-level collection and reporting of child well-being data was highlighted as an international priority by the OECD (OECD, 2021), identifying and incorporating instruments that capture PMH among children (11 years and under) in Canadian health surveys could help advance surveillance of PMH.

### Determinants

Our third and final research question pertained to the determinants of child PMH; two key conclusions can be drawn from the reviewed evidence. First, the results suggest that there is a wide range of factors that may be related to a child’s PMH. Second, while individual risk and protective factors were identified in the literature, there was an emphasis on the importance of socio-ecological factors influencing a child’s PMH. Indeed, many of the sources organized determinants based on different levels/domains. This is compatible with several existing mental health frameworks, including the Canadian conceptual framework for the youth and adult PMHSIF and the UK’s Conceptual Framework for Public Mental Health, which organize determinants within four socio-ecological levels (i.e., individual, family, community, society/structural) (Dykxhoorn et al., 2022; Orpana et al., 2016; PHAC, 2024). Similarly, Public Health Scotland organizes determinants of mental health for children and youth under five contextual constructs (i.e., individual, family, learning environment, community, structural) (Parkinson J, 2012). Given this, we decided to harmonize the presentation of determinants extracted from the scoping review by organizing them under the same four socio-ecological levels (i.e., individual, family, community, society) as Canada’s current PMHSIF for youth and adults (Orpana et al., 2016; PHAC, 2024).

At the individual level, healthy living behaviours (e.g., sleep, physical activity), overall physical health (e.g., physical condition, disability), and personal traits/identity (e.g., temperament, personal belief systems) were identified as potential factors related to child PMH. These determinants were not unexpected as similar risk and protective factors were also flagged as important during the development of Canada’s PMHSIF (Orpana et al., 2016). Play (CPHA, 2019; McLellan & Steward, 2015; Newland et al., 2015; Halton Region, 2023; UNICEF, 2018) and readiness for school (Aber & Jones, 1997; Child Trends, 2018) were two additional individual-level determinants pertinent to children that were flagged in this review. Play is key for a child’s development and has many benefits including learning critical thinking and social skills, fostering creativity, and ultimately enhancing physical and mental health (Ginsburg et al., 2007). Readiness for school encompasses a range of aspects such as motor skills, social and emotional capacities, and cognition (Ghandour et al., 2021; Gregory et al., 2021).

At the familial level, the importance of overall family dynamics, which includes relationships with parents/guardians, other family members (e.g., siblings), and even pets, was recognized. Relatedly, family socio-economic status, family structure (e.g., nuclear families, reconstituted families, lone-parents), home environment (e.g., adverse childhood experiences), and family health and health behaviours (e.g., parental substance misuse) were also identified as potential determinants for a child’s PMH. Many of these high-level determinants are consistent with the ones currently included in Canada’s PMHSIF (Orpana et al., 2016; PHAC, 2024). However, some of the more specific determinants in each category varied. For example, in the existing PMHSIF, the indicators for family relationships for youth focus on communication within the family generally and with their parents in particular, but indicators, such as relationships with other family members (e.g., siblings) and pets, are not currently included.

At the community level, school was flagged as a crucial setting for child PMH. School environment and experiences were also acknowledged as being an important determinant among youth in developing the PMHSIF (Orpana et al., 2016; PHAC, 2024). Some examples of school-related risk and protective factors that could influence a child’s PMH include engagement with learning, pressures and expectations, feelings of safety at school, and participation in school activities. Relationships with friends/peers (also key protective factors identified for youth and adults (Orpana et al., 2016; PHAC, 2024) as well as relationships with other adults (at school or in the community) were also highlighted. Other community-level (e.g., built and natural environment, connection to culture, neighbourhood factors) and society-level determinants (e.g., public policies, stigma/discrimination, housing conditions) that were extracted from this scoping review are represented in Canada’s PMHSIF (PHAC, 2024) or were considered as possible indicators during its development (Orpana et al., 2016).

Taken together, the evidence suggests that many of the determinants included in the conceptual framework for PMH in Canada (Orpana et al., 2016) may also be relevant for children. However, there are additional determinants that could be considered for inclusion in a child-specific PMHSIF. It is important to note that the determinants extracted from this scoping review were not all assessed similarly across sources. Indeed, some of the sources provided empirical support while others simply mentioned the determinants (without demonstrating its link to child PMH). Additional work will be needed to evaluate the relevance of some of these determinants prior to pursuing them for national surveillance. Moreover, some of the determinants missing from the existing PMHSIF (e.g., natural disasters, economic climate, family relations) that emerged in this review are not unique to childhood, and could be considered for inclusion in the conceptual PMH model for all populations (i.e., children, youth and adults).

### Strengths & Limitations

A key strength of this scoping review is that it synthesized evidence specific to PMH for children, which can be used to support the development of a child PMHSIF in Canada. Having routine national level reporting of child PMH could fill a knowledge gap in the Canadian context and address gaps identified internationally (OECD, 2021). Moreover, the findings from this scoping review could contribute to the development of child PMH indicators that could be added to existing MH surveillance systems in other Western countries. Another strength is the use of both peer-reviewed and grey literature sources, which allowed for the inclusion of over 100 sources containing various types of evidence. Lastly, the thematic analysis provided additional context and insight to the findings, facilitating connections between theories, concepts, and measures.

As with all scoping reviews of a broad topic such as child PMH, it is possible that certain sources were not captured in the search due to the keywords selected or search strategies. Another limitation is that the scoping review did not assess the quality of the evidence. Since information on theories/conceptual underpinnings that were not provided in the introduction/background section of a document (e.g., only mentioned in the discussion section to explain the findings) were not recorded, some may have been missed. This scoping review only included sources written in English and French, and conducted in Western countries, which could limit the generalizability of its findings internationally. Moreover, given the overrepresentation of sources from some countries and cultural differences across countries classified as Western, the findings may also vary in their generalizability to specific Western countries (Krys et al., 2022; Muthukrishna et al., 2020). This review synthesized literature on the PMH of children broadly and not for specific populations. While a few studies for specific groups such as Indigenous populations and refugee children were identified, information from those studies were included as part of the overall synthesis and not examined independently.

### Conclusion

Surveillance of child PMH is a gap in Canada’s current PMHSIF. This scoping review identified relevant theories, concepts/outcomes, instruments/measures, and determinants of PMH in children. Based on the findings, there does not appear to be significant conceptual differences in PMH for children compared to youth and adults. Many of the theories, concepts and outcomes, and determinants extracted for child PMH overlap with the existing PMH conceptual framework and PMHSIF in Canada. This suggests that the existing PMH conceptual framework and PMHSIF can be applied to children. However, slight modifications could be made to ensure child-relevant theories, instruments, and determinants are reflected. First, while many of the determinants identified in this review overlap with the existing PMHSIF, there are a few that are not currently included. Second, while the PMH concepts of hedonic and eudaimonic well-being are similar for children, youth, and adults, the way these concepts are measured can differ. It will be important to ensure that the instruments used to capture PMH among children are age-appropriate and validated. Third, to ensure that children’s perspectives/views are represented, a mix of both other-rated and self-rated instruments is recommended. Lastly, as only a few sources solely examined early childhood, most of the findings may be limited to middle childhood. Findings from this scoping review could be used to appraise the potential for adding child-specific indicators to the existing PMHSIF. Potential next steps could include 1) adapting the existing PMH conceptual framework (Orpana et al., 2016) to ensure that child-relevant concepts and determinants extracted from this review are reflected, 2) sharing the updated model with experts and partners/stakeholders for input, 3) searching available national health surveys for instruments of child PMH (including both instruments that were and were not flagged in this scoping review), and 4) conducting validation studies on identified instruments.

## Declarations

## Abbreviations

OECD: Organisation for Economic Co-operation and Development
PHAC: Public Health Agency of Canada
PMH: Positive mental health
PMHSIF: Positive Mental Health Surveillance Indicator Framework
WHO: World Health Organization

## Ethics approval and consent to participate

Not applicable.

## Availability of data and materials

All data generated or analysed during this study are included in this published article.

## Authors’ contributions

MV: Conceptualization, formal analysis, methodology, project administration, visualization, writing – original draft, writing – review & editing.

MJ: Formal analysis, methodology, validation, writing – review & editing.

CAC: Conceptualization, writing – review & editing. RD: Conceptualization, writing – review & editing.

LLO: Conceptualization, formal analysis, methodology, writing – review & editing.

## Acknowledgements

The authors would like to thank the research librarian Katherine Merucci (Health Canada) who helped with the search strategy for the grey literature and who gathered the peer-reviewed articles. We would like to extend our gratitude to Florence Lafontaine-Poissant (FLP) for her help in doing the screening (title/abstract and full-text) for the peer-reviewed literature. We would also like to thank Karen C. Roberts and Dana Paquette (Public Health Agency of Canada) for reviewing the manuscript.

The content and views expressed in this article are those of the authors and do not necessarily reflect those of the Government of Canada.

