## Appendices for "Positive mental health among children 11 years and under in Western countries: a scoping review to inform Canada’s public health surveillance"

(2) Statistics Canada, Ottawa, Ontario, Canada

**Caution: These appendices are for a preprint manuscript, which has not been peer-reviewed.**

Appendix 1. Peer-reviewed search strategy

Search Strategy

Medline

Database(s): Ovid MEDLINE(R) ALL 1946 to January 31, 2023
Search Strategy:

| **#** | **Searches** |
| --- | --- |
| 1 | exp *child/ or *adolescent/ or *minors/ or exp *infant/ |
| 2 | (((primary or elementary or middle or high or grade) adj3 school*) or (grade adj4 student*) or adolescen* or adrenarch* or babies or baby or boy? or child or children or childhood or daughter* or girl? or infan* or juvenile* or kid? or menarch* or neonat* or new born* or newborn* or paediatric* or pediatric* or perinatal or pre school* or preadolescen* or preadrenarch* or premenarch* or prepubert* or prepubescen* or preschool* or preteen* or pubert* or pubescen* or school age? or son or sons or stepchild* or stepdaughter* or stepson* or teen* or toddler* or young* or youth*).ti,kf. or (((primary or elementary or middle or high or grade) adj3 school*) or (grade adj4 student*) or adolescen* or adrenarch* or babies or baby or boy? or child or children or childhood or daughter* or girl? or infan* or juvenile* or kid? or menarch* or neonat* or new born* or newborn* or paediatric* or pediatric* or perinatal or pre school* or preadolescen* or preadrenarch* or premenarch* or prepubert* or prepubescen* or preschool* or preteen* or pubert* or pubescen* or school age? or son or sons or stepchild* or stepdaughter* or stepson* or teen* or toddler* or young* or youth*).ab. /freq=2 |
| 3 | or/1-2 [children] |
| 4 | health status indicators/ and mental health/ |
| 5 | ((indicator* or measure or measures or index or framework* or model or predictor? or metric? or criteria? or standard? or guideline? or measure* or indicator* or theory or theories or model? or framework* or best practice*) adj5 (mental* health* or emotion* or wellbeing or well-being)).ti,kf. |
| 6 | 4 or 5 |
| 7 | personal satisfaction/ or happiness/ |
| 8 | ((life or personal) adj3 satisf*).tw,kf. |
| 9 | (happy or happiness or enjoyment or enjoy* life or friendship? or healthy lifestyle?).tw,kf. |
| 10 | ((improv* or promot*) adj3 quality of life).tw,kf. |
| 11 | (positivity or (positive adj4 (mental health or emotional health or well being or wellbeing))).tw,kf. |
| 12 | or/7-11 |
| 13 | 3 and 6 and 12 |
| 14 | limit 13 to yr="2002-current" |
| 15 | 13 not 14 |

Embase

Database(s): Embase 1974 to 2023 January 31
Search Strategy:

| **#** | **Searches** |
| --- | --- |
| 1 | exp *child/ or *juvenile/ or *adolescent/ |
| 2 | (((primary or elementary or middle or high or grade) adj3 school*) or (grade adj4 student*) or adolescen* or adrenarch* or babies or baby or boy? or child or children or childhood or daughter* or girl? or infan* or juvenile* or kid? or menarch* or neonat* or new born* or newborn* or paediatric* or pediatric* or perinatal or pre school* or preadolescen* or preadrenarch* or premenarch* or prepubert* or prepubescen* or preschool* or preteen* or pubert* or pubescen* or school age? or son or sons or stepchild* or stepdaughter* or stepson* or teen* or toddler* or young* or youth*).ti,kf. or (((primary or elementary or middle or high or grade) adj3 school*) or (grade adj4 student*) or adolescen* or adrenarch* or babies or baby or boy? or child or children or childhood or daughter* or girl? or infan* or juvenile* or kid? or menarch* or neonat* or new born* or newborn* or paediatric* or pediatric* or perinatal or pre school* or preadolescen* or preadrenarch* or premenarch* or prepubert* or prepubescen* or preschool* or preteen* or pubert* or pubescen* or school age? or son or sons or stepchild* or stepdaughter* or stepson* or teen* or toddler* or young* or youth*).ab. /freq=2 |
| 3 | or/1-2 [children] |
| 4 | *health status indicator/ and exp *mental health/ |
| 5 | ((indicator* or measure or measures or index or framework* or model or predictor? or metric? or criteria? or standard? or guideline? or measure* or indicator* or theory or theories or model? or framework* or best practice*) adj5 (mental* health* or emotion* or wellbeing or well-being)).ti,kf. |
| 6 | 4 or 5 |
| 7 | Satisfaction/ or happiness/ |
| 8 | ((life or personal) adj3 satisf*).tw,kf. |
| 9 | (happy or happiness or enjoyment or enjoy* life or friendship? or healthy lifestyle?).tw,kf. |
| 10 | ((improv* or promot*) adj3 quality of life).tw,kf. |
| 11 | (positivity or (positive adj4 (mental health or emotional health or well being or wellbeing))).tw,kf. |
| 12 | or/7-11 |
| 13 | 3 and 6 and 12 |
| 14 | limit 13 to yr="2002-current" |
| 15 | 13 not 14 |
| 16 | limit 15 to (english or french) |

PsycINFO

Database(s): APA PsycInfo 1806 to January Week 4 2023
Search Strategy:

| **#** | **Searches** |
| --- | --- |
| 1 | adopted children/ or grandchildren/ or illegitimate children/ or stepchildren/ or only children/ or exp siblings/ or exp offspring/ or chronically ill children/ or childhood development/ or exp early childhood development/ or adolescent development/ or pediatrics/ or adolescent psychiatry/ or adolescent psychology/ or exp parent child relations/ or exp childrearing practices/ or exp child care/ or child custody/ or missing children/ or child psychiatry/ or child psychology/ |
| 2 | (((primary or elementary or middle or high or grade) adj3 school*) or (grade adj4 student*) or adolescen* or adrenarch* or babies or baby or boy? or child or children or childhood or daughter* or girl? or infan* or juvenile* or kid? or menarch* or neonat* or new born* or newborn* or paediatric* or pediatric* or perinatal or pre school* or preadolescen* or preadrenarch* or premenarch* or prepubert* or prepubescen* or preschool* or preteen* or pubert* or pubescen* or school age? or son or sons or stepchild* or stepdaughter* or stepson* or teen* or toddler* or young* or youth*).ti,id,hw. or (((primary or elementary or middle or high or grade) adj3 school*) or (grade adj4 student*) or adolescen* or adrenarch* or babies or baby or boy? or child or children or childhood or daughter* or girl? or infan* or juvenile* or kid? or menarch* or neonat* or new born* or newborn* or paediatric* or pediatric* or perinatal or pre school* or preadolescen* or preadrenarch* or premenarch* or prepubert* or prepubescen* or preschool* or preteen* or pubert* or pubescen* or school age? or son or sons or stepchild* or stepdaughter* or stepson* or teen* or toddler* or young* or youth*).ab. /freq=2 |
| 3 | or/1-2 [children] |
| 4 | ((indicator* or measure or measures or index or framework* or model or predictor? or metric? or criteria? or standard? or guideline? or measure* or indicator* or theory or theories or model? or framework* or best practice*) adj5 (mental* health* or emotion* or wellbeing or well-being)).ti,id,hw. |
| 5 | happiness/ |
| 6 | ((life or personal) adj3 satisf*).ti,ab,id,hw. |
| 7 | (happy or happiness or enjoyment or enjoy* life or friendship? or healthy lifestyle?).ti,ab,id,hw. |
| 8 | ((improv* or promot*) adj3 quality of life).ti,ab,id,hw. |
| 9 | (positivity or (positive adj4 (mental health or emotional health or well being or wellbeing))).ti,ab,id,hw. |
| 10 | or/5-9 |
| 11 | 3 and 4 and 10 |
| 12 | limit 11 to yr="2002-current" |
| 13 | 11 not 12 |
| 14 | limit 13 to (english or french) |

Scopus

( ( TITLE ( ( ( primary OR elementary OR middle OR high OR grade ) W/3 school* ) OR ( grade W/4 student* ) OR adolescen* OR adrenarch* OR babies OR baby OR boy* OR child OR children OR childhood OR daughter* OR girl* OR infan* OR juvenile* OR kid* OR menarch* OR neonat* OR "new born*" OR newborn* OR paediatric* OR pediatric* OR perinatal OR "pre school*" OR preadolescen* OR preadrenarch* OR premenarch* OR prepubert* OR prepubescen* OR preschool* OR preteen* OR pubert* OR pubescen* OR "school age*" OR son OR sons OR stepchild* OR stepdaughter* OR stepson* OR teen* OR toddler* OR young* OR youth* ) OR AUTHKEY ( ( ( primary OR elementary OR middle OR high OR grade ) W/3 school* ) OR ( grade W/4 student* ) OR adolescen* OR adrenarch* OR babies OR baby OR boy* OR child OR children OR childhood OR daughter* OR girl* OR infan* OR juvenile* OR kid* OR menarch* OR neonat* OR "new born*" OR newborn* OR paediatric* OR pediatric* OR perinatal OR "pre school*" OR preadolescen* OR preadrenarch* OR premenarch* OR prepubert* OR prepubescen* OR preschool* OR preteen* OR pubert* OR pubescen* OR "school age*" OR son OR sons OR stepchild* OR stepdaughter* OR stepson* OR teen* OR toddler* OR young* OR youth* ) ) ) AND ( TITLE ( ( indicator* OR measure OR measures OR index OR framework* OR model OR predictor* OR metric* OR criteria* OR standard* OR guideline* OR measure* OR indicator* OR theory OR theories OR model* OR framework* OR "best practice*" ) W/5 ( "mental* health*" OR emotion* OR wellbeing OR well-being ) ) ) AND ( TITLE-ABS-KEY ( ( life OR personal ) W/3 satisf* ) OR TITLE-ABS-KEY ( happy OR happiness OR enjoyment OR "enjoy* life" OR friendship* OR "healthy lifestyle*" ) OR TITLE-ABS-KEY ( ( improv* OR promot* ) W/3 "quality of life" ) OR TITLE-ABS-KEY ( positivity OR ( positive W/4 ( "mental health" OR "emotional health" OR "well being" OR wellbeing ) ) ) ) AND LANGUAGE ( english OR french )

Appendix 2. Grey literature search strategy

**DIRECT LINKS FROM RESEARCH LIBRARIAN**

Anderson Moore, Kristin; Lippman, Laura H.; McIntosh, Hugh (2009). Positive Indicators of Child Well-being: A conceptual framework, measures and methodological issues, Innocenti Working Papers, no. 2009-21, <https://www.unicef-irc.org/publications/580-positive-indicators-of-child-well-being-a-conceptual-framework-measures-and-methodological.html>

Child and Youth Health and Well-Being Indicators Project: CIHI and B.C. PHO Joint Summary Report <https://www2.gov.bc.ca/assets/gov/health/about-bc-s-health-care-system/office-of-the-provincial-health-officer/reports-publications/special-reports/child-health-2013.pdf>

Children and Young People's Mental Health and Wellbeing <https://fingertips.phe.org.uk/profile-group/mental-health/profile/cypmh>

Children's Bureau – Key Indicators of Child's Well-being <https://www.all4kids.org/news/blog/key-indicators-of-a-childs-well-being/>

Common Indicators of Social-Emotional Well-being in Early Childhood <https://www.childtrends.org/project/common-indicators-of-social-emotional-well-being-in-early-childhood>

Key national indicators of children’s health, development and wellbeing <https://www.aihw.gov.au/getmedia/366e81ce-eb90-470b-8464-a706c77d70f3/bulletin58.pdf.aspx?inline=true>

Measuring What Matters for Child Well-being and Policies <https://www.oecd.org/wise/measuring-what-matters-for-child-well-being-and-policies-e82fded1-en.htm>

Pan-Canadian Joint Consortium for School Health - Positive Mental Health Toolkit <http://www.jcsh-cces.ca/images/upload/JCSHPositiveMentalHealthToolkit.pdf>

Positive Mental Health Surveillance Indicator Framework <https://health-infobase.canada.ca/positive-mental-health/Publications>

Wisconsin - Child Well-Being Indicators Dashboard <https://children.wi.gov/Pages/ResearchData/Indicators.aspx>

**Grey Literature Search Strings**

| **Resource URL** | **Search String** |
| --- | --- |
| Google | child "mental health" indicator |
|  | (positive AROUND(4) mental health) children indicator |
|  | children positive mental health indicator |
|  | pediatric wellness indicator |
|  | enfants "santé mentale" indicateurs |
|  | enfants "Mieux-être" indicateurs |
|  | child* "mental health" indicator site:gov.uk |
|  | child* "mental health" indicator site:.ie |
|  | child* "mental health" indicator site:.au |
|  | child* "mental health" indicator site:.nz |
|  | enfants "santé mentale" indicateurs site:.fr |
| [Canada Commons](https://canadacommons.ca/search/?q=%28title%3A%28child*+OR+pediatric*+OR+paediatric*%29+AND+title%3A%28%22mental+health%22+OR+wellbeing+OR+%22well+being%22%29%29+AND+%28indicator*+OR+framework*+OR+criteria+OR+metric%29&i=items&advanced=yes&limit=50) | (title:(child* OR pediatric* OR paediatric*) AND title:("mental health" OR wellbeing OR "well being")) AND (indicator* OR framework* OR criteria OR metric) |
| [Canadian Research Institute for Law and the Family](https://prism.ucalgary.ca/handle/1880/107197) | "mental health" |
| [Science in French: Children's Health](https://hc-sc.libguides.com/c.php?g=725614&p=5198192) | Consult links in "healthy child development" box |

**Sources searched with no relevant results**

| **Resource URL** | **Search String** |
| --- | --- |
| JSTOR Open research reports <https://about.jstor.org/oa-and-free/open-research-reports/> | Child* "mental health" indicator* |
| Think tank search <https://guides.library.harvard.edu/hks/think_tank_search> | child* "mental health" positive |
|  | child* "mental health" |
| International government policy search <https://guides.library.utoronto.ca/ppg1000> | child* "mental health" |
| Intergovernmental organization search <https://guides.libraries.psu.edu/governmental-organizations> | child* "mental health" |
| Guidelines International Network <https://guidelines.ebmportal.com/> | "mental health" |
| MEDBOX – child health <https://www.medbox.org/categories#GO> | "mental health" |
| CAMH <https://www.camh.ca/> | Children indicators |
| Google | child "mental health" indicator site:.de |
